# Disrupted functional brain network associated with presence of hallucinations in Parkinson’s Disease

**DOI:** 10.1101/2024.09.30.24314594

**Authors:** Marcella Montagnese, Mitul A Mehta, Dominic ffytche, Michael Firbank, Rachael A Lawson, John-Paul Taylor, Edward T Bullmore, Sarah E Morgan

## Abstract

**Background:** Hallucinations negatively impact quality of life in Parkinson’s disease, yet their neural mechanisms remain poorly understood, particularly in early disease stages. This study aimed to identify functional connectivity differences associated with visual hallucinations in early Parkinson’s Disease and to validate these findings across independent datasets.

**Methods:** Resting state fMRI data from two prior independent studies was used (total N=185; N=84 hallucinators and N=101 non-hallucinators). Group differences in functional connectivity were assessed within predefined cytoarchitectonic cortical classes and functional networks, followed by whole-brain analysis using Network-Based Statistics (NBS). Associations with clinical measures, including hallucination severity, motor symptoms, cognition, and attention, were also evaluated.

**Results:** NBS identified a subnetwork of reduced functional connectivity in hallucinators, connecting regions involved in the default mode, somatomotor and attentional networks. This subnetwork was replicated in a matched sample from our independent cohort (N=50; p<0.01). Functional connectivity within the identified network was significantly associated with hallucination severity (R² = 0.35, p = 0.01), and with baseline and future motor symptoms, cognition, and attention in hallucinators.

**Conclusions:** The identified functional subnetwork shows promise as a potential biomarker and therapeutic target for Parkinson’s disease psychosis, warranting further investigation and validation in future studies.

## Introduction

Psychosis is a common yet underappreciated feature of Parkinson’s Disease (PD), with estimates suggesting half of the 10 million PD patients worldwide experience psychosis at some point during their illness^1^. Symptoms include hallucinations, predominantly visual, which have been shown to negatively affect patients’ and carers’ quality of life and predict dementia^2,3^. However, the mechanisms underlying visual hallucinations (VH) and their relationship to cognitive processing and clinical outcomes remain poorly understood. VH might be underpinned not just by regions with neural pathology, but also by unaffected regions within broader functional networks^4–6^. Converging evidence suggests the contribution of attentional network dysfunctions and an imbalance in top-down and bottom-up perceptual processing^7^, implicating multiple regions and functional networks^5,6,8–12^.

Against this backdrop, two main models of PD Psychosis (PDP) emerge as particularly relevant: the Perception-Attention-Deficit (PAD)^13^ and the Attentional Network Dysfunction^14^ model. The PAD model proposes that patients experience deficits in sensory activation and attentional binding. Hallucinations arise from erroneous sensory activation of inappropriate proto-objects (early visual processing where sensory inputs compete for recognition as distinct objects in visual awareness) and failures in attentional binding. The Attentional Network Dysfunction model also attributes VH to attentional control deficits, specifically from perturbations in the interactions among Dorsal Attention Network (DAN), Ventral Attention Network (VAN), and Default Mode Network (DMN) - with experimental work supporting this^15,16^.

A systems-level analysis of functional network connections could support or challenge these models. Nonetheless, to date, little work has investigated whole-brain resting-state functional connectivity (FC) in early-stage PDP. Previous studies have examined psychosis in established disease, primarily due to recruitment challenges^2^, and most studies have used task-based fMRI, investigating changes in PD participants with visual hallucinations (PDVH) during visual tasks of varying complexity^16–19^. While these studies shed light on functional, cognitive, and visual alterations in PDVH, the variability in tasks makes replication difficult. Moreover, VH are linked to deficits across multiple cognitive domains^9,20^. In this context, resting-state fMRI is a valuable complementary approach to examine whole-brain FC across domains^21,22^. Lastly, while some studies have used seed-based-analyses of resting-state fMRI or looked at circumscribed networks^12,23–26^, few have performed whole-brain, data-driven analyses^9,10^.

The overall aim and hypotheses of this study were therefore to:

A) Investigate functional connectivity markers in PD patients with hallucinations by:

(i) Evaluating group differences in global connectivity. Based on prior research^9^, we hypothesized reduced global FC in patients compared to healthy controls, with more pronounced reductions in PDVH than PDNOVH.
(ii) Comparing group differences across Yeo functional networks and von Economo classes. We expected lower FC in attentional networks (DAN,VAN) and higher FC in the Default Mode Network (DMN) in PDVH.
(iii) Using Network Based Statistics (NBS) to identify brain-wide connectivity differences associated with VH in PD.
B) Assess whether any significant results from A) replicated in an independent cohort.
C) Explore the relationships between hallucination-specific network differences and clinical/cognitive variables, both cross-sectionally and longitudinally.

## Methods and Materials

### Primary cohort-PPMI

Our primary sample came from the PPMI (Parkinson’s Progression Marker Initiative) and included resting state fMRI data from PD patients and age-matched healthy controls (HC). Detailed exclusion criteria for the PPMI cohort are given in **Supplementary Section 4**. Patients were grouped into visual hallucination (PDVH) or non-visual hallucination (PDNOVH) groups based on scoring ≥1 on Question 1.2 of the Movement Disorder Society Unified Parkinson’s Disease Rating Scale (MDS-UPDRS^27^) - ‘*Over the past week have you seen, heard, smelled or felt things that were not really there?*’ - at one or more concomitant/previous assessments (N=12 scored 1, N=11 scored 2, and N=2 scored 3). The overall sample included: *N*=25 PDVH patients, *N*=56 PDNOVH patients and *N*=24 healthy controls (HC) ^28^. By design, most PPMI patients were at early stages of their disease.

### Cognitive measures

Global cognition was assessed with the Montreal Cognitive Assessment (MoCA)^29^; visuospatial function with the Benton Judgement of Line Orientation task^30^; and executive function with semantic fluency (total number of animals named in one minute)^31^. Attention was measured with the Letter-number sequencing task^32^, and episodic memory with the Hopkins Verbal Learning Test Total Delayed Recall^33^ (sum of Recall Trial 1-3).

### Clinical measures

Motor symptom severity was measured using the motor subscale of total MDS-UPDRS-III^27^. PD duration was recorded as number of years since PD diagnosis. Levodopa-equivalent daily dose (LEDD) was the daily dose (mg) on assessment day (See PPMI manual^34^). Sleep disorders were assessed with the REM Sleep Behaviour Disorder Screening Questionnaire (RBDSQ)^35^. Cerebrospinal fluid (CSF) sampling and analysis is described in detail in the PPMI manual^34^. The following were available for a subset of patients: CSF β-amyloid (Aβ1– 42 in pg/mL), tau proteins total (t-Tau) and α-synuclein concentration levels.

### Secondary Cohort - ICICLE-PD

Our replication cohort came from the ICICLE-PD study^36^. Inclusion and exclusion criteria are given in **Supplementary Section 4**. Most participants completed the North East Visual Hallucination Interview (NEVHI)^30^, a semi-structured interview developed by Mosimann et al^37^ covering the phenomenology of VH. Patient and caregiver versions were used. A positive score (“Yes”) on one or more of NEVHI Part A screening questions 1.1 to 1.6 about presence of hallucinatory experiences (such as Question 1.1 “*Do you feel like your eyes ever play tricks on you? Have you ever seen something (or things) that other people could not see*?”) was used to categorise patients as PDVH or PDNOVH (see **Supplementary Section 5**). To replicate findings from the PPMI cohort, two subsamples were created from ICICLE-PD: Sample 1 (N=96) included patients that were well-matched to each other in clinical and demographic variables; Sample 2 (N=50) included a subset of patients matched to both each other and to the PPMI cohort based on key variables (age, sex, disease severity (MDS-UPDRS-III), cognition (MoCA), and years of education). Further details regarding the matching process and cohort characteristics can be found in the **Results**, **Table 1**, **Supplementary Table 1,** and **Supplementary Section 4**.

**Table 1.**
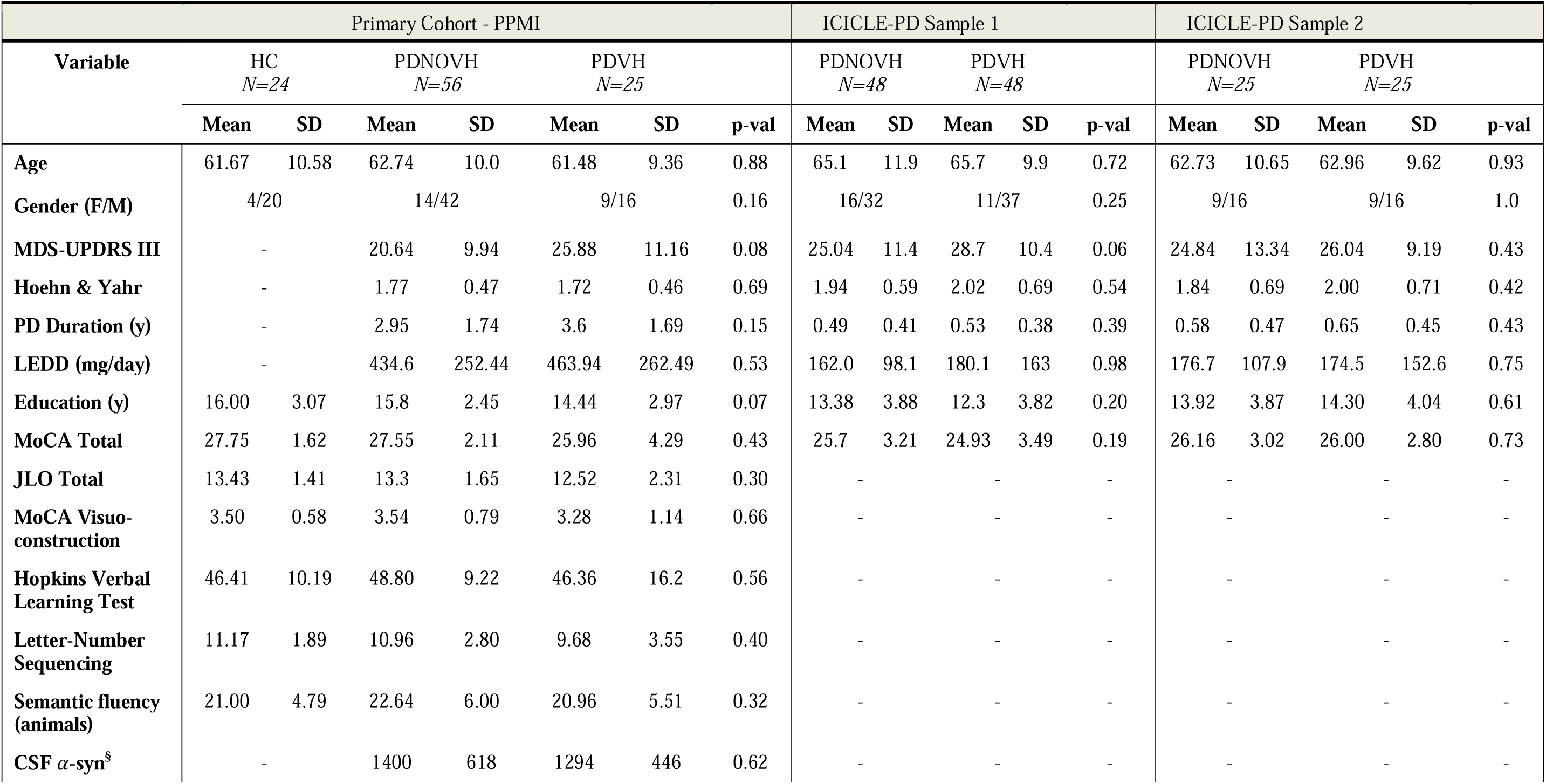
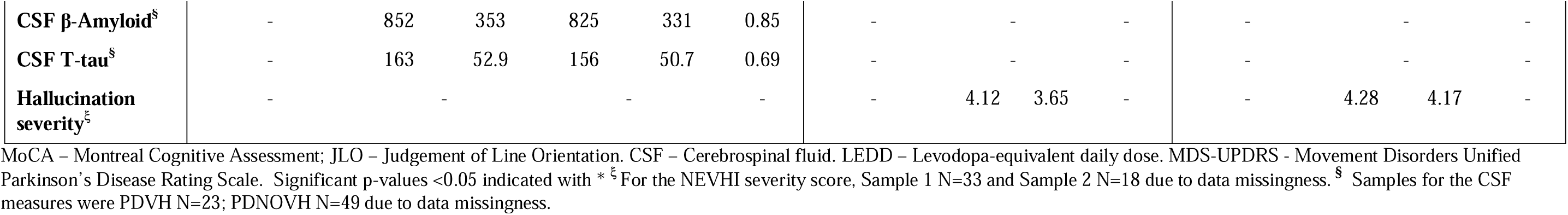
Summary demographics for the primary and secondary cohorts. , including the group of matched ICICLE-PD patients included in the NBS replication analysis.

### Clinical and Cognitive measures

Hallucinations severity was computed using the NEVHI by following the guidelines outlined in its manual^37^. This involved multiplying scores of hallucination frequency, hallucination duration, and severity factors specific for the type of hallucination (see **Supplementary Section 6)**. As for the PPMI cohort, global cognition was assessed with the Montreal Cognitive Assessment (MoCA)^29^ and motor symptom severity was measured using the motor subscale of total MDS-UPDRS-III^27^.

### Statistical analysis of demographics

Demographic and clinical characteristics of PDVH, PDNOVH and HC groups were compared using one-way ANOVA for normally distributed and Kruskall-Wallis for non-normally distributed variables, with χ^2^ for categorical variables. For variables only relevant to patients, independent t-tests and Mann-Whitney tests were used. Shapiro-Wilk tests were used to assess normality.

### Data Pre-processing

Full details on the acquisition of both MPRAGE (Magnetization Prepared Rapid Acquisition Gradient Echo) T1-weighted sequence and resting state fMRI data in PPMI and ICICLE-PD can be found in **Supplementary Section 1**. rsfMRI data for both cohorts was pre-processed according to a published pipeline^38^ described in **Supplementary Section 2**. Denoising of motion artefacts was done with BrainWavelet toolbox’s wavelet despiking^38^. Brain surfaces were parcellated using an atlas with 308 cortical regions of approximately equal size based on a subdivision of the Desikan–Killiany atlas^39^. FC matrices were calculated using Pearson’s correlation of pairwise normalized wavelet coefficients between regions.

Participants with mean framewise displacement (FD) >0.6 mm were excluded to reduce motion artefacts^40^: *N*=4 PDVH, *N*=8 PDNOVH and *N*=1 HC were excluded in PPMI; and *N*=3 PDNOVH and *N*=5 PDVH in ICICLE-PD. Mean FD was regressed from each FC matrix edge to remove motion correlations and distance-dependent motion effects (**Supplementary Section 3)**. In ICICLE-PD, further 9 patients (4 PDVH, 5 PDNOVH) were excluded due to failed co-registration, and 1 PDNOVH was excluded for having many drop-out regions.

### Functional Connectivity analyses across Yeo networks and von Economo classes in PPMI

We first calculated group differences in mean FC averaged across the whole brain and averaged within each von Economo cytoarchitectonic class and Yeo network. Differences were calculated between the three groups using linear models (including age, sex and age*sex as covariates). To assess sensitivity, analyses were repeated including MoCA, UPDRS-III and LEDD as covariates. After removing outliers (±2 SDs), sample sizes were: PDVH N=22, PDNOVH N=55, HC N= 22.

### Network-based statistics (NBS) in PPMI

We used Network Based Statistics (NBS)^41^ to identify a subnetwork of edges that differed between the PDVH and PDNOVH groups, co-varying for sex and age. NBS analyses were run with 5,000 permutations to ascribe a FWER-corrected p-value (<0.05). All analyses were performed using the Network-Based Statistics Toolbox *v1.2*^41^.

### NBS replication analysis in ICICLE-PD Cohort

We tested whether the NBS subnetwork identified using the PPMI cohort replicated in the ICICLE-PD cohort. Replication was assessed using two approaches: 1) Calculating total FC within the NBS network for each individual in the ICICLE-PD cohort and applying a linear model to test group differences, controlling for age and sex; and 2) using the NBS toolbox to directly compare PDVH and PDNOVH group differences in ICICLE-PD, using the same t-threshold and covariates (age and sex) as in PPMI.

### Relating NBS connectivity to clinical and cognitive variables

#### Primary PPMI cohort

To understand the relationship between the NBS network defined in PPMI and clinical and cognitive variables of interest we ran a Principal Component Analysis (PCA) on all clinical and cognitive variables available with R’s *factoextra* and *factominer* (*v2.4*). We then related the PCA components to average NBS subnetwork connectivity with Spearman’s correlations - run separately for PDVH and PDNOVH groups.

Further exploratory Spearman correlation analyses were conducted to assess whether average NBS connectivity was associated with cognitive decline and clinical outcomes at baseline and at follow-up. To maximize data availability, a follow-up period of 4 years (± 1 year) was used. The variables included in the analysis were attentional performance, MoCA, RBD, and MDS-UPDRS-III scores.

#### Secondary ICICLE-PD cohort

In the PDVH group from Sample 2, we related average FC within the PPMI NBS network to hallucination severity scores from the NEVHI. A linear regression was run with masked NBS FC as the independent variable and severity as the dependent variable, with a sensitivity analysis controlling for disease severity (MDS-UPDRS III).

## Results

### Primary Sample (PPMI) Characteristics

There were no statistically significant group differences across shared demographic and cognitive variables, nor in medication and motor symptoms severity between PDVH and PDNOVH; see **Table 1**.

### Replication Sample (ICICLE-PD) Characteristics

In the full ICICLE-PD cohort (N=104), the PDVH and PDNOVH groups were unmatched on key demographics and clinical variables (sex, disease severity, MoCA – see **Supplementary Table 1**). To address this, we matched the groups on all shared clinical and demographic variables, creating **Sample 1** (N=48 PDVH, N=48 PDNOVH). However, patients in Sample 1 were still not well-matched to the PPMI cohort, with the ICICLE-PD ones having fewer years of education, lower LEDD, and shorter disease duration (all *p*<0.05, see **Supplementary Section 4**). Sample 2 was therefore introduced (*N*=25 PDVH, *N*=25 PDNOVH), where ICICLE-PD patients were matched both with each other *and* with PPMI. While disease duration and dopaminergic medication still differed between ICICLE-PD and PPMI, Sample 2 was otherwise well-matched to PPMI on age, sex, disease severity (MDS UPDRS-III), cognition (total MoCA) and years of education (see **Table 1** and **Supplementary Section 4**).

### Group differences in Functional Connectivity in PPMI cohort

In the PPMI cohort, FC decreased across groups with highest values in HC and lowest values in PDVH (see **Figure 1A**), however, differences were not statistically significant (*Kruskal-Wallis test-statistic*=2.65, *p*=0.266).

**Figure 1.**
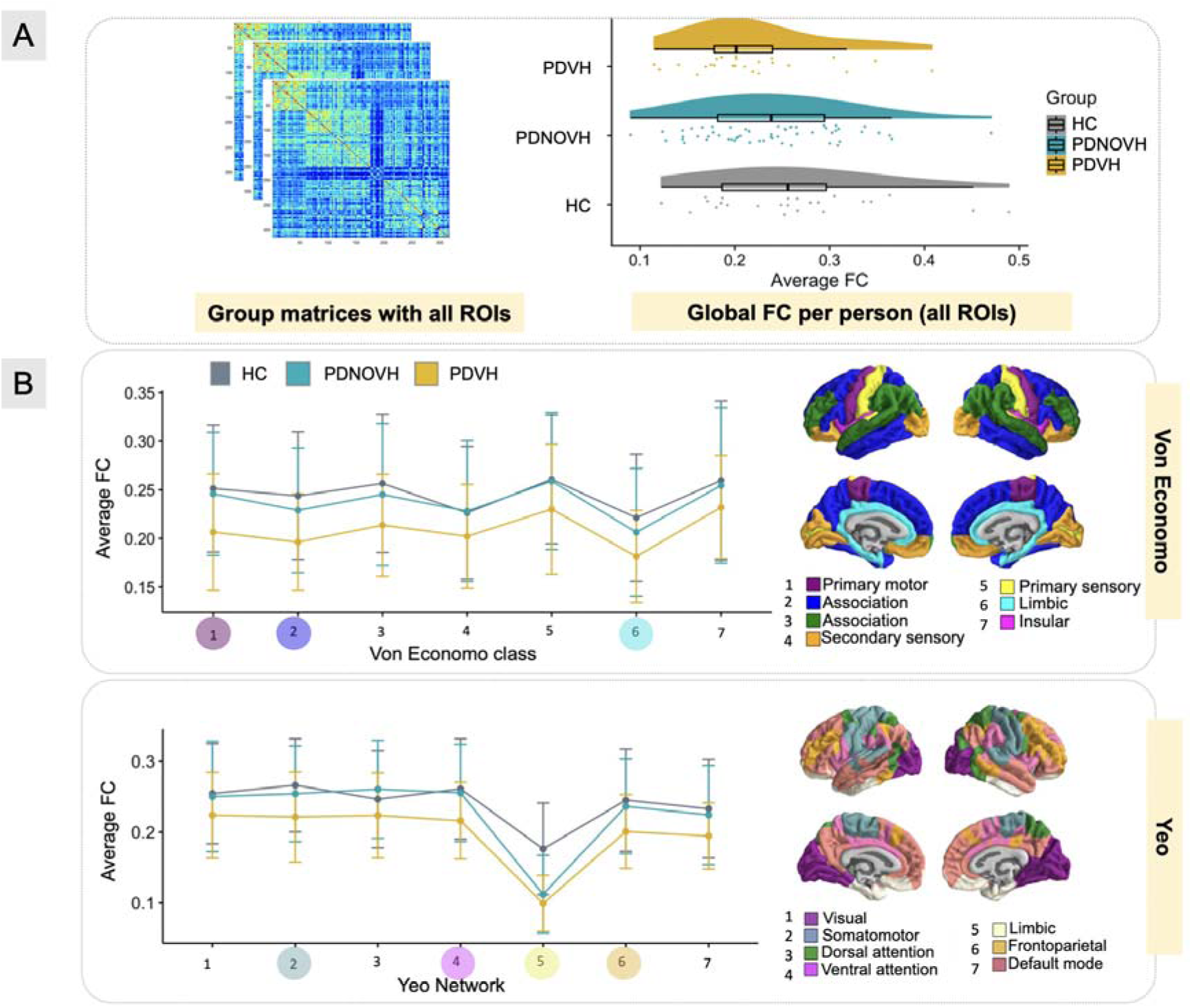
Group differences in functional connectivity (FC) at the global level and within previously defined von Economo and Yeo functional networks. (A) Left: average FC matrix for each group. Right: differences in global FC across the three groups. (B) Average FC within all seven von Economo cytoarchitectonic classes and seven Yeo canonical functional networks. Error bars indicate standard deviation. Coloured dots in (B) show the networks and classes where a significant group effect was found (p<0.05) *before* False Discovery Rate (FDR) correction and where the same colour indicates the relevant class or network.

Group comparisons of FC across functional Yeo networks and von Economo cytoarchitectonic classes (**Figure 1B**) showed differences across all groups for the primary motor (*p*=0.02), first association (*p*=0.01) and limbic (*p*=0.04) von Economo classes, alongside somatomotor (*p*=0.02), ventral attention (*p*=0.03), limbic (*p*<0.001) and frontoparietal Yeo networks (*p*=0.037). For these classes and networks, post-hoc tests revealed that PDVH had significantly lower mean FC than HC and PDNOVH. However, after False Discovery Rate (FDR) correction no result remained significant, except for the functional Yeo limbic network (*p_FDR_* = 0.007) where both PD groups differed significantly from HC. **Supplementary Table 2**.

### NBS subnetwork related to hallucinations in PPMI cohort

NBS analyses revealed a statistically significant network of reduced FC in PDVH compared to PDNOVH (**Figure 2**) comprising 22 nodes and 23 edges (t-threshold=3.8, *p*=0.04, 5,000 permutations). Connections were mainly between regions located in the DMN, somatomotor, DAN and VAN networks (see **Figure 2C** and **Supplementary Table 3)**. The edge-pairs with the highest t-statistics were between the DMN and DAN, DMN and somatomotor, and DAN and somatomotor networks. No significant NBS network was found in the opposite direction (PDVH FC > PDNOVH).

**Figure 2.**
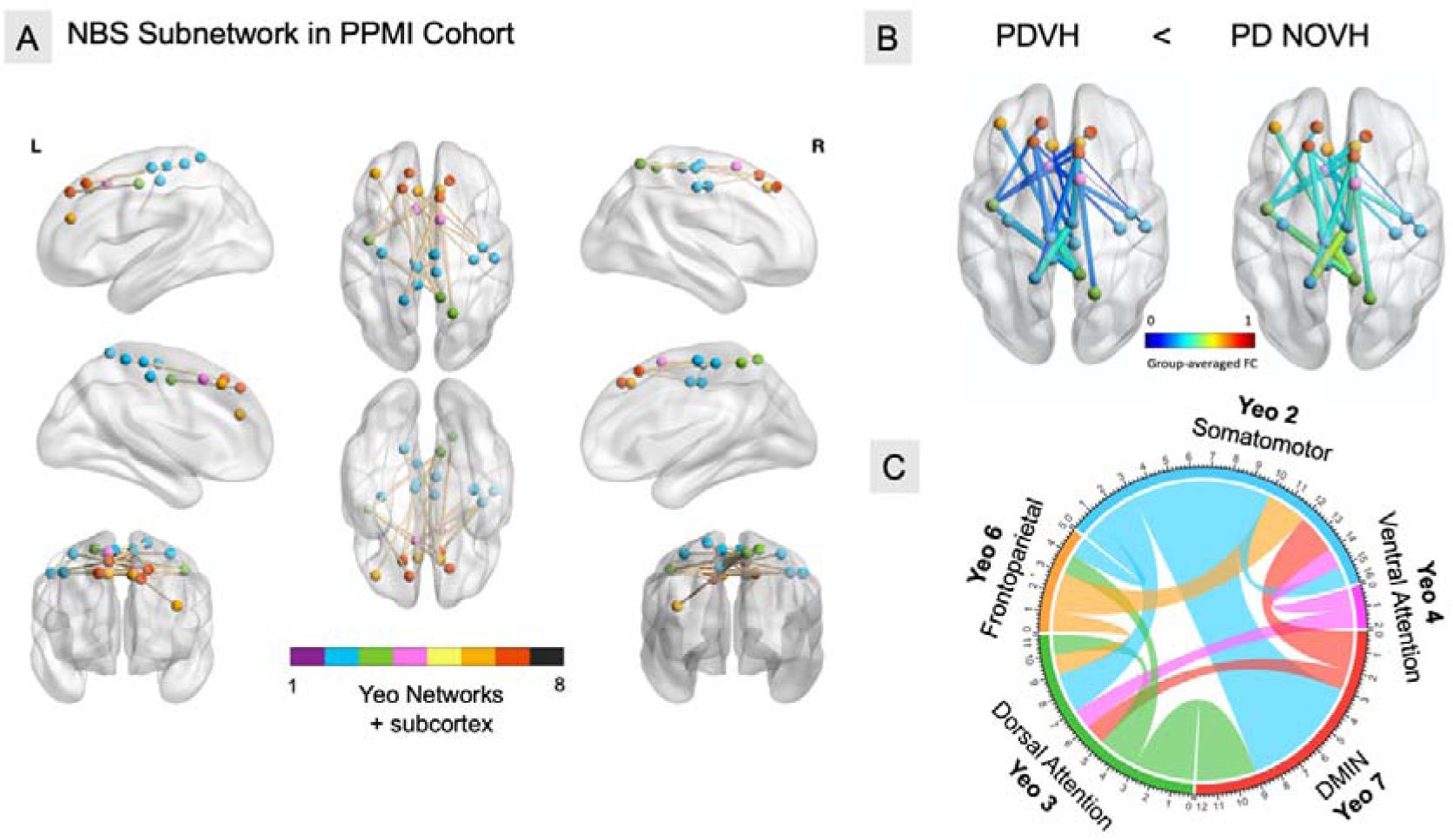
Network Based Statistics differences in functional connectivity between the VH and NOVH groups, in the PPMI Cohort. Nodes are colour-coded according to their mapping to the 7 canonical functional networks published by Yeo *et al.* (2011). (A) Network of edges showing reduced FC in the VH group compared to the NOVH group, identified by the NBS analysis. An overall network of 22 nodes and 23 edges was identified by testing a one-tailed hypothesis. (B) The same NBS network shown in part (A), with edges coloured according to group averaged FC. (C) Connectogram visualisation of the NBS network, highlighting the most salient Yeo networks.

### Replication of NBS subnetwork in ICICLE-PD cohort

We assessed whether the PPMI NBS subnetwork replicated in an independent dataset. For ICICLE-PD Sample 1, there was no significant difference in FC within the PPMI-defined NBS network between the PDVH and PDNOVH groups (*t-test statistic*=0.2, *p*>0.05). When using NBS to calculate PDVH vs PDNOVH connectivity differences directly in Sample 1, we found no significant NBS network (*threshold*= 3.8, contrast PDVH<PDNOVH, age and sex as covariates).

We then turned to Sample 2 – where ICICLE-PD patients were matched not only to each other but also to the PPMI cohort for variables including years of education and MoCA scores. We observed a significant reduction in FC within the PPMI-defined NBS network in PDVH compared to PDNOVH (*t-test statistics* 7.31, *p*<0.01) - **Figure 3C**. We then re-ran the NBS analysis with the same parameters used in PPMI (*t-threshold*=3.8, 5,000 permutations, contrast PDVH<PDNOVH, age and sex as covariates) and again identified a significant network (*p*=0.03) of lower FC in patients with hallucinations vs. without hallucinations. This was larger (44 nodes, 46 edges) than the PPMI NBS network and contained almost the entire original PPMI NBS network (17/23 original edges), see **Figure 3B** and **Supplementary Table 4**.

**Figure 3.**
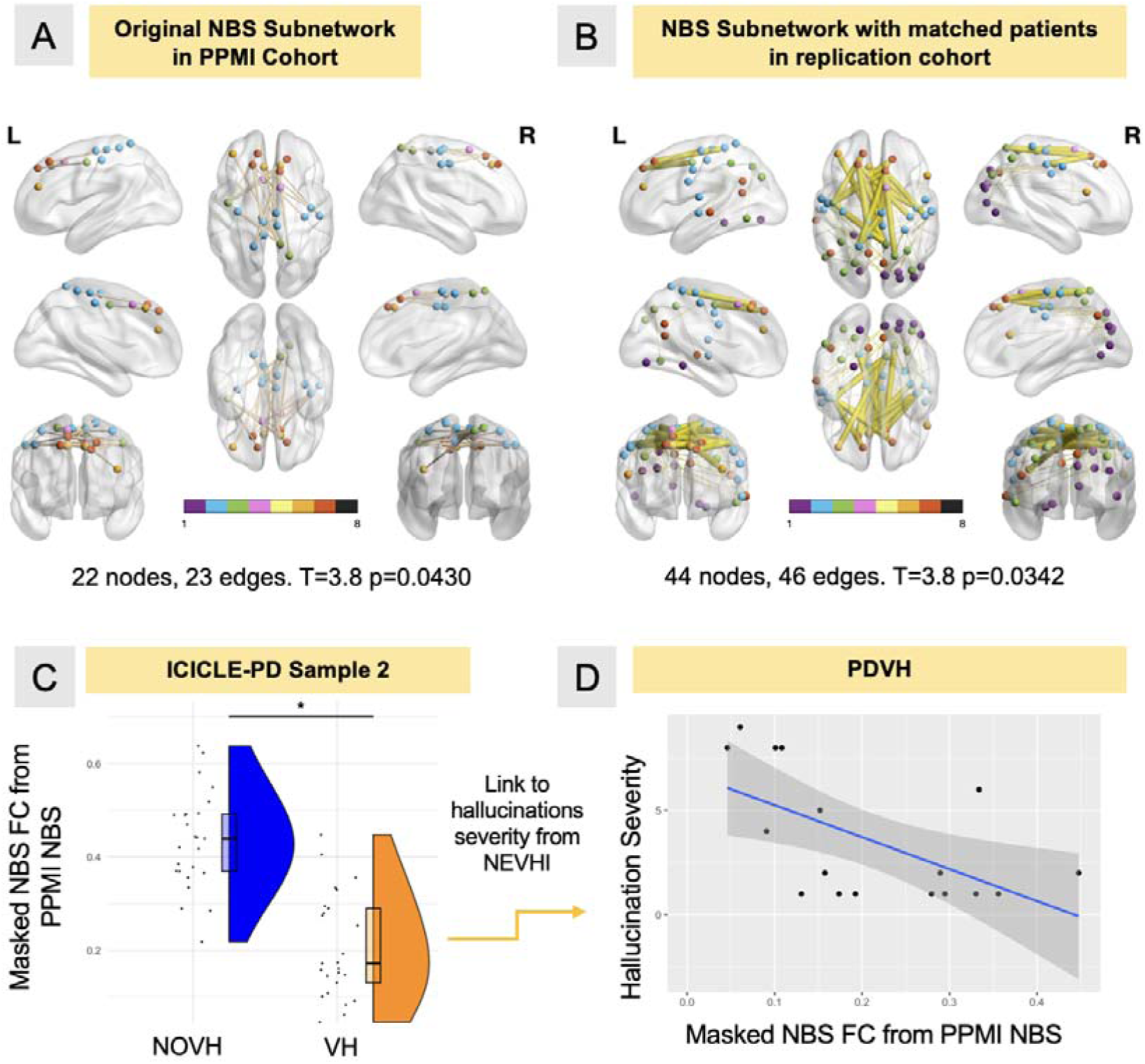
NBS network replicated in the ICICLE-PD dataset. A) Original NBS subnetwork from the PPMI cohort. B) NBS subnetwork calculated from matched patients in replication cohort (VH vs NOVH) where PDVH have lower FC than NOVH. Overlapping edges with the PPMI result in part A) are coloured yellow. Nodes are colour-coded according to their mapping to the 7 canonical functional networks by Yeo *et al.* (2011). C) Raincloud plot of FC within the PPMI-defined NBS subnetwork in both groups of the ICICLE-PD cohort – with boxes showing data quartiles, whiskers indicating the full data range, dots indicating single-individual values, and half-violin plots depicting the density of the data. D) Negative correlation between composite hallucination severity score and FC within the (PPMI-defined) NBS subnetwork in the ICICLE-PD VH group (N=17).

FC within the PPMI-defined NBS subnetwork was significantly associated with hallucination severity in the PDVH group from ICICLE-PD Sample 2 (*R*^2^ = 0.35, *adjusted R*^2^ = 0.31, *p*=0.01; **Figure 3D)**. This remained significant (*R*^2^ = 0.36, *adjusted R*^2^ = 0.27, *p*=0.01) when controlling for MDS-UPDRS III.

### Relation between NBS connectivity and clinical/cognitive measures (PPMI cohort)

We related FC within our NBS subnetwork to clinical and cognitive measures from the PPMI cohort. (N=69, including N=23 PDVH, N=46 PDNOVH, after excluding cases with missing CSF data). Principal Component Analysis (PCA) identified 4 dimensions with eigenvalues >1, explaining 63.03% of total variance. High PC1 was associated with lower age and better attention, executive function and general cognition (MoCA), and to a lesser extent, lower motor severity. High PC2 was related to higher CSF levels (alpha-synuclein, t-tau, beta-amyloid). High PC3 was related to longer PD duration, higher LEDD, worse RBD symptoms and better visuospatial processing. High PC4 was related to higher LEDD, better memory, and older age (**see Figure 4**).

**Figure 4.**
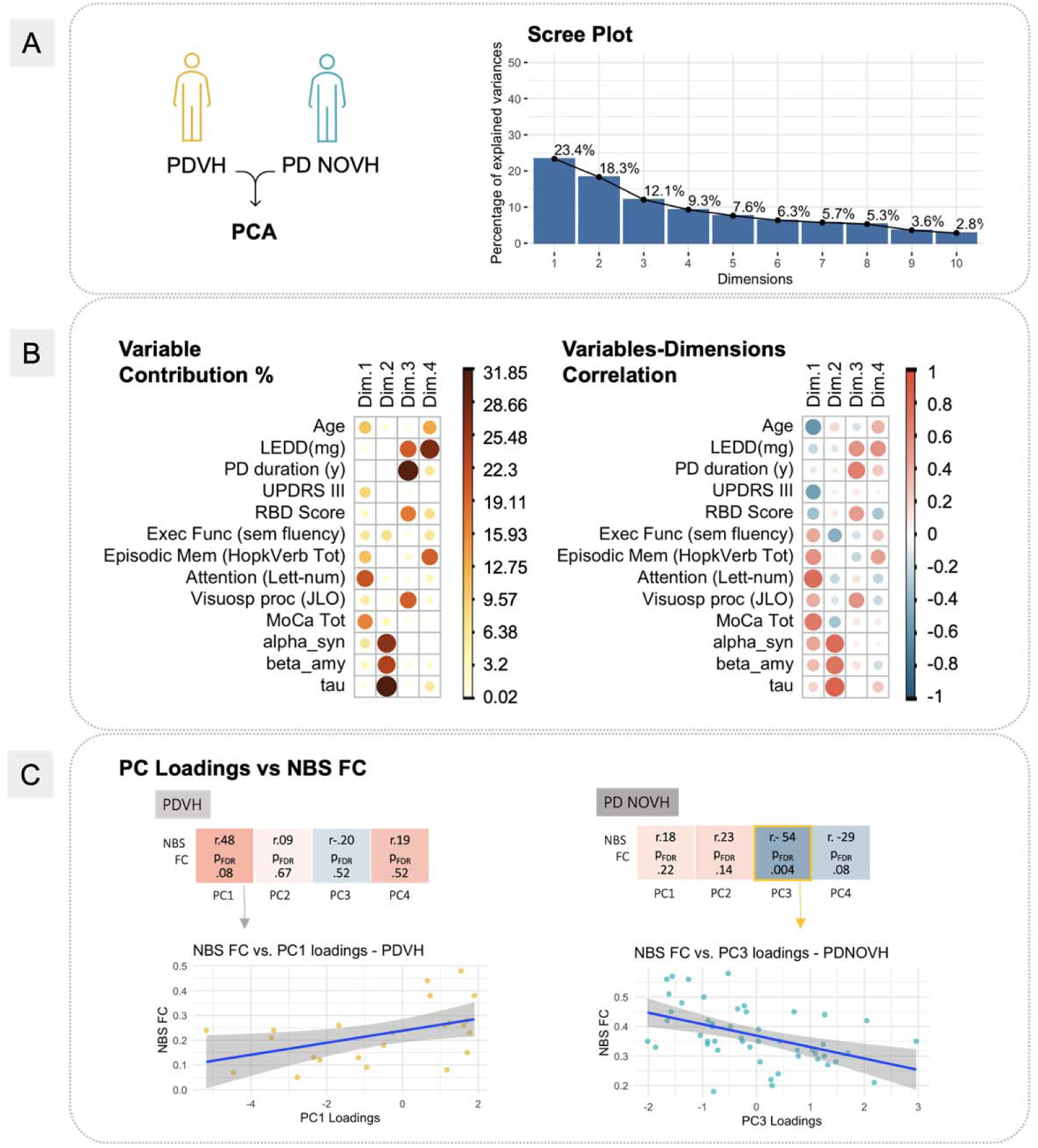
Principal Component Analysis (PCA) of clinical and cognitive variables, for the PPMI dataset. A) Scree Plot showing the amount of variance explained by the PCs. Only components with eigenvalues >1 were selected, resulting in 4 principal components. B) On the left, visual representation of the first 4 dimensions with a plot of the relative contribution of each variable (in %) to each of the dimensions. The colours represent the contributing weights with dark red in both plots indicating a higher contribution (%). On the right, the correlation between each variable and each dimension. C) Spearman’s correlation analyses linking NBS FC to each of the 4 PCs for each group separately. On the bottom left, we show a correlation plot in the PDVH group for NBS FC vs the loading of the PC1 dimension; on the bottom right we show a correlation plot in the PDNOVH group for NBS FC vs the loading of the statistically significant PC3 dimension.

In PDVH, connectivity in the NBS subnetwork positively correlated with PC1, but the relationship was not significant after FDR correction (ρ=0.48, *p-val*=0.02, *p-val_FDR_*= 0.08).

In PDNOVH, NBS FC was negatively correlated with PC3 both before and after FDR correction (ρ= -0.54, *p-val*=0.001, *p-val_FDR_*= 0.004), indicating that lower NBS FC was associated with higher LEDD, longer disease duration, worse RBD symptoms, and better visuospatial processing.

### Relation between NBS connectivity and future clinical outcomes in PPMI Cohort

Lastly, we examined the association between baseline NBS connectivity and future clinical outcomes (see **Figure 5**). In the PDVH group, average NBS FC was positively correlated with better attention at both baseline and follow-up, and better cognition (MoCA total score) at follow-up. Lower NBS FC at baseline were related to worse (higher) MDS-UPDRS III scores at follow-up. No significant correlations were found for the PDNOVH group.

**Figure 5.**
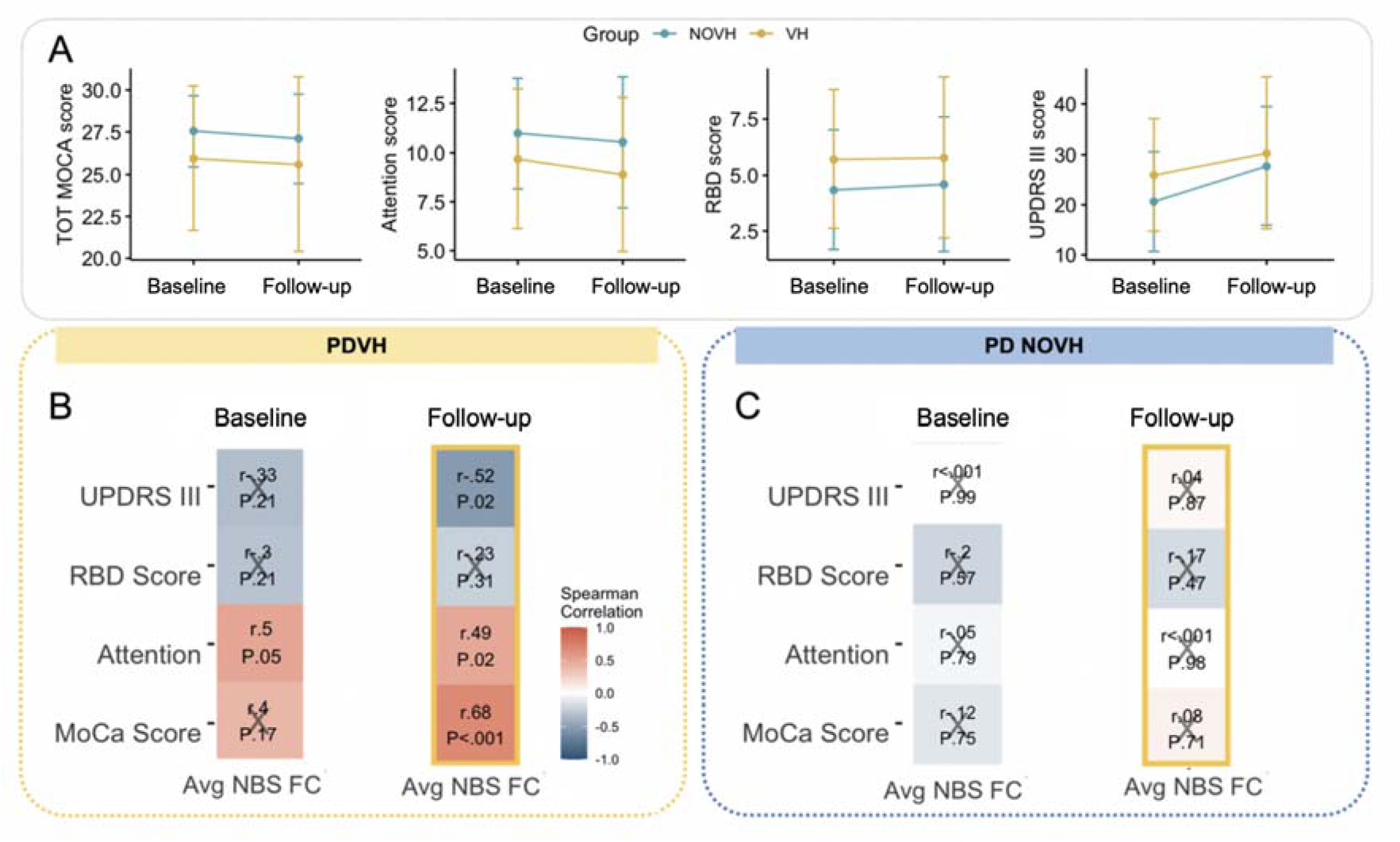
Relation between NBS connectivity and future clinical outcomes. A) Plot of mean and SD for each variable per group at baseline and at follow-up time of 4 years (± 1 year). B) Correlations between clinical measures and FC within the NBS subnetwork at baseline and at follow-up, for the PDVH group. Spearman correlation coefficients are given, alongside p-values after FDR correction. C) The same correlations for the PDNOVH group.

## Discussion

NBS identified a replicable network of reduced functional connectivity associated with visual hallucinations in PD patients. The NBS network included connections within and across the dorsal attention, ventral attention, and default mode networks, and replicated in an independent dataset when cohorts were matched for clinical and demographic variables. Lower functional connectivity within the network was associated with increased hallucination severity, and with baseline and future scores of motor symptom severity, cognition, and attention in PD patients with hallucinations.

### Relation to prior models of PD Psychosis

The involvement of the DAN, VAN and DMN in the NBS network of reduced functional connectivity in PD psychosis was in-line with prior work^6,8,9,16,24,42,43^. Several network edges connected the DAN with the DMN, the somatomotor network and the frontoparietal network, in-keeping with the central role of the DAN in PD psychosis^10,24,42^ and consistent with both the Attentional deficit^14^ and the Perception-Attention-Deficit (PAD)^13^ models. Further underscoring the notion of attentional impairments in PD hallucinators, many of the NBS-identified nodes map onto anatomical landmarks within the superior frontal gyrus— a principal component of the DAN and a structure implicated in PD psychosis^8,9^.

Our results suggest a complex and nuanced interpretation of the DMN’s role in PD hallucinations. Contrary to one of our hypotheses (ii) and to previous work^16,26,44^, our results did not reveal hyperconnectivity within the DMN in PDVH, nor between the DMN and other networks. A partial explanation for this discrepancy is that, to our knowledge, no previous studies had examined the DMN with the same whole-brain analytic approach that we employed. Another possible explanation is that studies reporting hyperactivity of the DMN often included older patients with longer disease duration: e.g. in Shine et al.^16^ PDVH patients had mean age of 69.3 years and disease duration of 6 years (compared to 61.4 and 3.6 years, respectively in the PPMI cohort). Similarly in Yao et al^26^ the mean age of PDVH patients was 67.6 years old, with an average 10 years disease duration.

### Validation and relation to hallucination severity

The NBS network replicated in an independent cohort (ICICLE-PD) matched to the original cohort in terms of age, sex, disease severity, education, and cognition. Prior to matching, the secondary cohort had lower cognitive scores and years in education, suggesting the NBS network may characterize hallucinators with relatively preserved cognition or higher education. This is in-keeping with the hypothesis that differences in cognitive performance introduce heterogeneity into the network characteristics of different patient groups^20^. Our NBS network might also represent a characteristic neural signature of PD hallucinators at an early disease stage, when major cognitive decline has not yet occurred, but initial dysfunction of attentional networks has started to emerge. At present the different effects of age, education, cognition, and disease duration on FC differences are difficult to disentangle, but these could be further explored in future work with better phenotyped longitudinal cohorts.

In the replication cohort, more severe hallucinations were related to reduced NBS FC, establishing an important, direct link between NBS network connectivity and degree of hallucinations. We note that higher severity composite score could mean that lower NBS FC is linked to patients experiencing multiple types of hallucinations or the same symptoms occurring more frequently or lasting longer in time. More work is required to delineate these options^45^.

### Cognitive significance

The relationships between NBS network connectivity and clinical/cognitive measures aligned with attentional deficits in PD psychosis, corroborating prior meta-analytic findings of attention, executive function, and general cognition being most impaired in hallucinators^20^. Finally, higher NBS connectivity at baseline correlated both with better performance on attentional tasks and overall cognition, and with worsening motor symptoms severity at follow-up. The specificity of these predictions to the PDVH group suggests again that the NBS network is characteristic of PD psychosis.

#### Limitations

Some limitations should be noted: (i) participants were not recruited to investigate psychotic symptoms, resulting in a lack of detailed phenotyping about hallucination characteristics. Consequently, reliance on MDS-UPDRS question 1.2 may have included patients with minor and/or complex hallucinations, including experiences beyond the visual domain. Future studies should incorporate more comprehensive measures across sensory modalities^46^; (ii) visual testing was not conducted to assess potential low-level visual deficits^47^, and hallucination states could not be directly measured during scanning, limiting the ability to examine connections with active hallucinatory episodes versus only hallucination traits; (iii) we lacked comprehensive data on non-anti-Parkinsonian drugs that could induce hallucinations (e.g., anticholinergics, opioids). While LEDD was included as a covariate, other medications may have influenced the results; (iv) sample sizes, whilst comparable to other recent studies, were relatively small. Nonetheless, the NBS approach helped maximize statistical power, and replication of the NBS network in an independent dataset matched for key variables is promising.

#### Conclusions

In conclusion, we identified a replicable network of reduced functional connectivity associated with PD psychosis. The network included regions from key attentional and default mode networks and its association with hallucination severity, as well as with baseline and future motor symptoms, cognition, and attention, underscores its relevance in Parkinson’s Disease psychosis. Overall, these results highlight the importance of resting-state fMRI network analysis in uncovering replicable neural differences related to hallucinations, and a deeper understanding of early-stage brain network dysfunction in Parkinson’s Disease psychosis could help target therapeutic interventions toward the most effective neural systems.

## Abbreviations

PDP: Parkinson’s Disease Psychosis;
VH: Visual Hallucinations;
FC: Functional Connectivity;
PDVH: Parkinson’s Disease with Hallucinations;
PDNOVH: Parkinson’s Disease without Hallucinations;
PPMI: Parkinson’s Progression Markers Initiative;
ICICLE: Incidence of Cognitive Impairments in Cohorts with Longitudinal Evaluation.

## Data Availability

All analyses used publicly available packages and code, available at: https://github.com/marcellamontagnese/PDPrestingfMRI. No new data was generated. PPMI data, downloaded on 05/10/2021, can be accessed at www.ppmi-info.org, and ICICLE-PD data is available by contacting the study lead at Newcastle University (http://bam-ncl.co.uk/our-work/studies/icicle-pd/).

## Acknowledgements

We thank Dr Daniel Williams for the valuable discussion during the revision of this work. The overall funding bodies for each of the authors are as follows: Marcella Montagnese was supported during her PhD work by the NIHR Maudsley Biomedical Research Centre. This paper represents independent research funded by the National Institute for Health Research (NIHR) Biomedical Research Centre at South London and Maudsley NHS Foundation Trust and King’s College London. The views expressed are those of the authors and not necessarily those of the NHS, the NIHR or the Department of Health and Social Care.

J-P. Taylor and M. Firbank are supported by the NIHR Newcastle Biomedical Research Centre (BRC) based at Newcastle upon Tyne Hospitals NHS Foundation Trust and Newcastle University. RA Lawson is supported by a Janet Owens Parkinson’s UK Senior Research Fellowship (F-1801). M. A. Mehta is funded by the Research England. His current research is supported by grants from the Medical Research Council (UK), Alzheimers Research UK, NIHR, Wellcome Trust, Lundbeck and SoseiHeptares. D. ffytche was funded by the NIHR Programme Grants for Applied Research (SHAPED: RP-PG-0610-10100). E. Bullmore was supported by an NIHR Senior Investigator award and the Wellcome Trust collaborative award for the Neuroscience in Psychiatry Network. S.E. Morgan was supported by the Accelerate Programme for Scientific Discovery, funded by Schmidt Futures.

Data were curated and analysed using a computational facility provided by King’s College London.

In addition to the listed authors, we thank the following members of the ICICLE-PD Study Group who all made a significant contribution to the work reported in this paper: Roger Barker (John van Geest Centre for Brain Repair, University of Cambridge, UK, Principal Investigator), Patrick F Chinnery (Institute of Genetic Medicine, Newcastle University, UK, Principal Investigator); John T O’Brien (Department of Psychiatry, University of Cambridge, UK, Principal Investigator); Trevor W Robbins (Department of Psychology, University of Cambridge, UK, Principal Investigator); Gordon Duncan and David Breen (Anne Rowling Regenerative Neurology Clinic, University of Edinburgh, UK, Site Investigators), Caroline H Williams-Gray, Marta Camach, Gemma A Cummins, Jonathan Evans, Ruwani Wijeyekoon, Kirsten Scott, Tom Stoker, Julia Greenland, Natalie Valle Guzman, Lucy Collins, Simon Stott and Sarah Mason (John van Geest Centre for Brain Repair, University of Cambridge, UK, Site Investigators); David J Brooks (Department of Medicine, Imperial College, London, UK, Principal Investigator); Tien K Khoo (School of Medicine and Menzies Health Institute Queensland, Griffith University, Australia, Site Investigator), Fionnuala Johnston and Claire McDonald (Translational and Clinical Research Institute, UK, Site Investigator); James B Rowe (Behavioural and Clinical Neuroscience Institute, UK, Site Investigator); David J Burn and Lynn Rochester (Translational and Clinical Research Institute, Newcastle University, Newcastle Upon Tyne, UK, Site Investigators);

The views expressed are those of the authors and not necessarily those of the NIH, NHS, the NIHR or the Department of Health and Social Care.

ICICLE-PD was funded by Parkinson’s UK (J-0802, G-1301, G-1507). The research was supported by the Lockhart Parkinson’s Disease Research Fund, National Institute for Health Research (NIHR) Newcastle Biomedical Research Unit and Centre based at Newcastle upon Tyne Hospitals NHS Foundation Trust and Newcastle University and the NIHR Cambridge Biomedical Research Centre (NIHR203312).

## Authors’ Contributions

M.Montagnese: conceptualization, methodology, formal analysis, writing (original draft preparation, review and editing); S.E.Morgan: conceptualization, methodology, supervision, writing (original draft preparation, review and editing); M.Mehta, D.ffytche: supervision, conceptualization, investigation, methodology, writing (review and editing); M.Firbank, R.Lawson, JP.Taylor: dataset access, writing (review and editing).

## Supplementary material

Supplementary material for this paper is attached.

## References

1. Fenelon G. Hallucinations in Parkinson’s disease: Prevalence, phenomenology and risk factors. Brain. 2000;123(4):733–745. doi:10.1093/brain/123.4.733

2. ffytche DH, Creese B, Politis M, et al. The psychosis spectrum in Parkinson disease. Nat Publ Group. 2017;13. doi:10.1038/nrneurol.2016.200

3. ffytche DH, Aarsland D. Psychosis in Parkinson’s Disease. Vol 133. (Chaudhuri KR, Titova N, eds.). Academic Press Inc.; 2017. doi:10.1016/bs.irn.2017.04.005

4. ffytche DH. The hodology of hallucinations. Cortex. 2008;44(8):1067–1083. doi:10.1016/J.CORTEX.2008.04.005

5. Collerton D, Barnes J, Diederich NJ, et al. Understanding visual hallucinations: a new synthesis. Neurosci Biobehav Rev. Published online May 2, 2023:105208. doi:10.1016/j.neubiorev.2023.105208

6. Pagonabarraga J, Bejr-Kasem H, Martinez-Horta S, Kulisevsky J. Parkinson disease psychosis: from phenomenology to neurobiological mechanisms. Nat Rev Neurol. Published online January 15, 2024:1–16. doi:10.1038/s41582-023-00918-8

7. Zarkali A, Adams RA, Psarras S, Leyland LA, Rees G, Weil RS. Increased weighting on prior knowledge in Lewy body-associated visual hallucinations. Brain Commun. 2019;1(1):fcz007. doi:10.1093/braincomms/fcz007

8. Muller AJ, Shine JM, Halliday GM, Lewis SJG. Visual hallucinations in Parkinson’s disease: Theoretical models. Mov Disord. 2014;29(13):1591–1598. doi:10.1002/mds.26004

9. Hepp DH, Foncke EMJ, Dubbelink KTEO, Berg WDJVD, Berendse HW, Schoonheim MM. Loss of functional connectivity in patients with Parkinson disease and visual hallucinations. Radiology. 2017;285(3):896–903. doi:10.1148/radiol.2017170438

10. Tan JB, Müller EJ, Orlando IF, et al. Abnormal higher-order network interactions in Parkinson’s disease visual hallucinations. Brain. Published online September 7, 2023:awad305. doi:10.1093/brain/awad305

11. O’callaghan C, Hall JM, Tomassini A, et al. Visual Hallucinations Are Characterized by Impaired Sensory Evidence Accumulation: Insights From Hierarchical Drift Diffusion Modeling in Parkinson’s Disease. Biol Psychiatry Cogn Neurosci Neuroimaging. 2017;2:680–688. doi:10.1016/j.bpsc.2017.04.007

12. Thomas GEC, Zeidman P, Sultana T, Zarkali A, Razi A, Weil RS. Changes in both top-down and bottom-up effective connectivity drive visual hallucinations in Parkinson’s disease. Brain Commun. 2023;5(1):fcac329. doi:10.1093/braincomms/fcac329

13. Collerton D, Perry E, McKeith I. Why people see things that are not there: a novel perception and attention deficit model for recurrent complex visual hallucinations. Behav Brain Sci. 2005;28(6):737–757.

14. Shine JM, Halliday GM, Naismith SL, Lewis SJG. Visual misperceptions and hallucinations in Parkinson’s disease: Dysfunction of attentional control networks? Mov Disord. 2011;26(12):2154–2159. doi:10.1002/mds.23896

15. Shine JM, Halliday GM, Gilat M, et al. The role of dysfunctional attentional control networks in visual misperceptions in Parkinson’s disease. Hum Brain Mapp. 2014;35(5):2206–2219. doi:10.1002/hbm.22321

16. Shine JM, Muller AJ, O’Callaghan C, Hornberger M, Halliday GM, Lewis SJ. Abnormal connectivity between the default mode and the visual system underlies the manifestation of visual hallucinations in Parkinson’s disease: a task-based fMRI study. Npj Park Dis. 2015;1(1):1–8. doi:10.1038/npjparkd.2015.3

17. Stebbins GT, Goetz GG, Carrillo MC, et al. Altered cortical visual processing in PD with hallucinations: An fMRI study. Neurology. 2004;63(8 PG-1409-1416):1409–1416. doi:10.1212/01.WNL.0000141853.27081.BD

18. Ramirez-Ruiz B, Marti MJ, Tolosa E, et al. Brain Response to Complex Visual Stimuli in Parkinson’s Patients with Hallucinations: A Functional Magnetic Resonance Imaging Study. Mov Disord. 2008;23(16 PG-2335-2343):2335–2343. doi:10.1002/mds.22258

19. Meppelink AM, De Jong BM, Renken R, Leenders KL, Cornelissen FW, Van Laar T. Impaired visual processing preceding image recognition in Parkinson’s disease patients with visual hallucinations. Brain. 2009;132(11 PG-2980-2993):2980–2993. doi:10.1093/brain/awp223

20. Montagnese M, Vignando M, ffytche D, Mehta MA. Cognitive and visual processing performance in Parkinson’s disease patients with vs without visual hallucinations: A meta-analysis. Cortex. 2022;146:161–172. doi:10.1016/j.cortex.2021.11.001

21. Bijsterbosch J, Smith SM, Beckmann CF. Introduction to Resting State fMRI Functional Connectivity. Oxford University Press; 2017.

22. Chou Y hui, Sundman M, Whitson HE, et al. Maintenance and Representation of Mind Wandering during Resting-State fMRI. Sci Rep. 2017;7(1):40722. doi:10.1038/srep40722

23. Bejr-kasem H, Pagonabarraga J, Martínez-Horta S, et al. Disruption of the default mode network and its intrinsic functional connectivity underlies minor hallucinations in Parkinson’s disease. Mov Disord. 2019;34(1):78–86. doi:10.1002/mds.27557

24. Walpola IC, Muller AJ, Hall JM, et al. Mind-wandering in Parkinson’s disease hallucinations reflects primary visual and default network coupling. Cortex. 2020;125:233–245. doi:10.1016/j.cortex.2019.12.023

25. Yao N, Pang S, Cheung C, et al. Resting activity in visual and corticostriatal pathways in Parkinson’s disease with hallucinations. Parkinsonism Relat Disord. 2015;21(2 PG-131-137):131–137. doi:10.1016/j.parkreldis.2014.11.020

26. Yao N, Shek-Kwan Chang R, Cheung C, et al. The default mode network is disrupted in parkinson’s disease with visual hallucinations. Hum Brain Mapp. 2014;35(11 PG-5658-5666):5658–5666. doi:10.1002/hbm.22577

27. Goetz CG, Tilley BC, Shaftman SR, et al. Movement Disorder Society-sponsored revision of the Unified Parkinson’s Disease Rating Scale (MDS-UPDRS): scale presentation and clinimetric testing results. Mov Disord Off J Mov Disord Soc. 2008;23(15):2129–2170. doi:10.1002/mds.22340

28. Marek K, Jennings D, Lasch S, et al. The Parkinson Progression Marker Initiative (PPMI). Prog Neurobiol. 2011;95:629–635. doi:10.1016/j.pneurobio.2011.09.005

29. Nasreddine ZS, Phillips NA, Bédirian V, et al. The Montreal Cognitive Assessment, MoCA: A Brief Screening Tool For Mild Cognitive Impairment. J Am Geriatr Soc. 2005;53(4):695–699. doi:10.1111/j.1532-5415.2005.53221.x

30. Benton AL, Varney NR, Hamsher KD. Visuospatial judgment. A clinical test. Arch Neurol. 1978;35(6):364–367. doi:10.1001/archneur.1978.00500300038006

31. Ardila A, OstroskylJSolís F, Bernal B. Cognitive testing toward the future: The example of Semantic Verbal Fluency (ANIMALS). Int J Psychol. 2006;41(5):324–332. doi:10.1080/00207590500345542

32. Pezzuti L, Rossetti S. Letter-Number Sequencing, Figure Weights, and Cancellation subtests of WAIS-IV administered to elders. Personal Individ Differ. 2017;104:352–356. doi:10.1016/j.paid.2016.08.019

33. Benedict RHB, Schretlen D, Groninger L, Brandt J. Hopkins Verbal Learning Test— Revised: Normative data and analysis of inter-form and test–retest reliability. Clin Neuropsychol. 1998;12(1):43–55. doi:10.1076/clin.12.1.43.1726

34. Parkinson’s Progression Markers Initiative Research Documents and SOPs |. Accessed June 26, 2023. https://www.ppmi-info.org/study-design/research-documents-and-sops

35. Stiasny-Kolster K, Mayer G, Schäfer S, Möller JC, Heinzel-Gutenbrunner M, Oertel WH. The REM sleep behavior disorder screening questionnaire—A new diagnostic instrument. Mov Disord. 2007;22(16):2386–2393. doi:10.1002/mds.21740

36. Yarnall AJ, Breen DP, Duncan GW, et al. Characterizing mild cognitive impairment in incident Parkinson disease: The ICICLE-PD Study. Neurology. 2014;82(4):308–316. doi:10.1212/WNL.0000000000000066

37. Mosimann UP, Collerton D, Dudley R, et al. A semi-structured interview to assess visual hallucinations in older people. Int J Geriatr Psychiatry. 2008;23(7):712–718. doi:10.1002/gps.1965

38. Patel AX, Kundu P, Rubinov M, et al. A wavelet method for modeling and despiking motion artifacts from resting-state fMRI time series. NeuroImage. 2014;95:287–304. doi:10.1016/j.neuroimage.2014.03.012

39. Romero-Garcia R, Atienza M, Clemmensen LH, Cantero JL. Effects of network resolution on topological properties of human neocortex. NeuroImage. 2012;59(4):3522–3532. doi:10.1016/j.neuroimage.2011.10.086

40. Váša F, Bullmore ET, Patel AX. Probabilistic thresholding of functional connectomes: Application to schizophrenia. NeuroImage. 2018;172:326–340. doi:10.1016/j.neuroimage.2017.12.043

41. Zalesky A, Fornito A, Bullmore ET. Network-based statistic: Identifying differences in brain networks. NeuroImage. 2010;53(4):1197–1207. doi:10.1016/j.neuroimage.2010.06.041

42. Shine JM, Keogh R, O’Callaghan C, Muller AJ, Lewis SJG, Pearson J. Imagine that: Elevated sensory strength of mental imagery in individuals with Parkinson’s disease and visual hallucinations. Proc R Soc B Biol Sci. 2015;282(1798):20142047. doi:10.1098/rspb.2014.2047

43. Bhome R, Thomas GEC, Zarkali A, Weil RS. Structural and Functional Imaging Correlates of Visual Hallucinations in Parkinson’s Disease. Curr Neurol Neurosci Rep. 2023;23(6):287–299. doi:10.1007/s11910-023-01267-1

44. Baggio HC, Segura B, Junque C. Resting-State Functional Brain Networks in Parkinson’s Disease. CNS Neurosci Ther. 2015;21(10):793–801. doi:10.1111/cns.12417

45. Montagnese M, Vignando M, Collerton D, et al. Cognition, hallucination severity and hallucination-specific insight in neurodegenerative disorders and eye disease. Cognit Neuropsychiatry. 2021;27(2-3):105–121. doi:10.1080/13546805.2021.1960812

46. Montagnese M, Leptourgos P, Fernyhough C, et al. A Review of Multimodal Hallucinations: Categorization, Assessment, Theoretical Perspectives, and Clinical Recommendations. Schizophr Bull. Published online August 9, 2020. doi:10.1093/schbul/sbaa101

47. Weil R, Schrag AE, Warren JD, Crutch SJ, Lees AJ, Morris HR. Visual dysfunction in Parkinson’s disease. Brain. 2016;139(11):2827–2843. doi:10.1093/brain/aww175

