## Supplementary Material for "Disrupted functional brain network associated with presence of hallucinations in Parkinson’s Disease"

1. **Resting and structural MRI Data Acquisition**

**PPMI**

All T1 MRI data was acquired using a MPRAGE (magnetization prepared rapid acquisition gradient echo) 3D T1-weighted sequence on 3T scanners. The acquisition parameters were as follows: sagittal plane, voxel size 1 × 1 × 1 mm, repetition time TR = 2300 ms, echo time TE = 3 ms, flip angle = 9°, 256x256x240 mm matrix. Acquisition time was ∼ 5 minutes.
At selected sites, resting state functional MRI (rsfMRI) was also performed with participants keeping their eyes open. Images were acquired with a gradient-echo imaging sequence lasting for ∼ 8.5 minutes with the following parameters: repetition time TR = 2400 ms, echo time TE = 25 ms, flip angle = 80°, slices in the ascending direction, slice thickness = 3.3 mm, voxel size 3.3 × 3.3 × 3.3 mm^3^, 210 volumes.

### ICICLE-PD All T1 MRI data was acquired using a MPRAGE T1-weighted sequence on 3T scanners with an eight-channel receiver head coil. The acquisition parameters were as follows: sagittal plane, voxel size 1 × 1 × 1 mm, TR = 2250 ms, TE = 2.98 ms, flip angle = 9°, 256x256x196 mm matrix. Acquisition time was shorter than for the PPMI cohort (< 5 minutes). Assessments were completed in an ‘on’ motor state. rsfMRI was also completed by all patients included in the current work. The echo-planar imaging sequence was run on 3T scanners with the following parameters: repetition time TR = 3000 ms, echo time TE = 40 ms, flip angle = 90°, sequential slices in the ascending order, voxel size 3.25 × 3.25 × 6 mm^3^.

### 2. Detailed information about the pre-processing pipeline

Overall, the following were performed using a combination of FreeSurfer, AFNI, FSL, Python and Matlab on a high-performance Cluster:

- Manual inspection of T1-weighted images to ensure no large motion artefacts were present.
- Running the recon-all command in FreeSurfer for cortical reconstruction, skull-stripping, segmentation of cortical grey and white matter and reconstruction of the cortical surface and grey-white matter boundary (Fischl et al., 1999).
- Running the speedyPPX script (Patel et al., 2014) for slice timing correction, rigid-body head movement correction, obliquity transform of the rsfMRI images to the T1w ones, affine co-registration of the rsfMRI images to the structural ones using a grey-matter mask. Images were mapped to the MNI152 standard space with non-linear warping. No bandpass filter was applied up to this point.
- Denoising motion artefacts using the time-series data. This was achieved with the wavelet despiking method from the BrainWavelet toolbox ([www.brainwavelet.org](http://www.brainwavelet.org), (Patel et al., 2014). Briefly, this method uses a wavelet decomposition of the signal to correct for head movements. The main advantage of this unsupervised method is that it is able to remove several types of spatially heterogeneous and non-linear movement-induced artefacts (e.g., slower ones like spin-history effects, as well as high-frequency ones like spikes) by detecting non-stationary events across different frequencies (Patel et al., 2014).
- Manual check of the co-registrations for all participants and that the wavelet despiking worked.
- Extraction of mean motion parameters:
  - mean framewise displacement (FD) – calculated as the sum of the absolute derivatives of the 6 motion parameters (x, y, z, α, β, γ), representing 3 planes of translation and 3 planes of rotation. The mean was then taken to obtain one value per subject.
  - mean spike percentage (SP) – as described by Patel et al., (2014) this is defined as “the percentage of grey matter voxels containing a spike in that frame of data” at any given point. The mean was calculated as in the step above.
- Brain parcellation using the APARC500 and Glasser atlases (Glasser et al., 2016). Drop-out regions were also calculated. The APARC500 is an atlas developed by (Romero-Garcia et al., 2012) and is based on a subdivision of the Desikan-Killiany atlas (Desikan et al., 2006). The APARC500 comprises of 308 cortical ROIs that are spatially contiguous and of approximately equal size (∼5mm) generated by using a backtracking algorithm described in detail in the original publication.
- Bandpass wavelets filter the data at scale 2 (corresponding to 0.02–0.1 Hz) in accordance with our TR values. Correlation matrices were calculated with the Pearson’s correlation coefficient by correlating pairwise the (normalised) wavelet coefficients between regions. This resulted in subject-specific 308 × 308 cortical symmetric FC matrices and 324 x 324 cortical + subcortical matrices. FC matrices were cut-down depending on how many drop-out regions were present. Overall, the final FC matrices included 316 ROIs, including subcortical regions. One PDVH patient was excluded due to a significantly shorter acquisition scan and higher number of drop-out regions.
- Participants with mean FD above 0.6 mm were excluded as done in previous work (Váša et al., 2018) to ensure minimal motion artefacts. Based on this and on quality check of the resulting timeseries, 4 PDVH and 8 PD NOVH patients were excluded, as well as 1 HC participant. The final samples included N=25 PDVH, N=56 PD NOVH and N=24 HC.

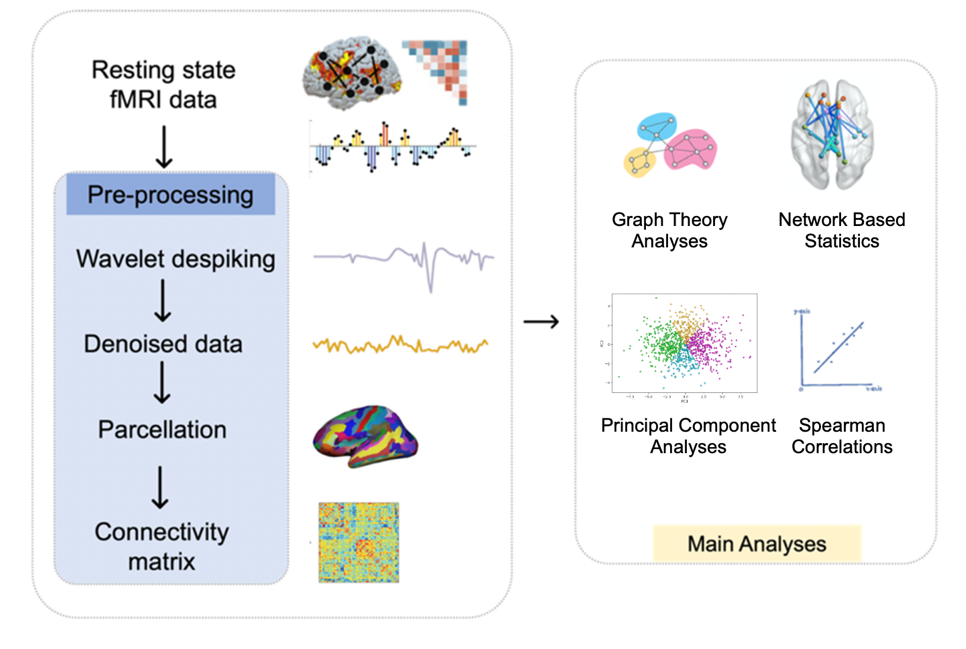

**Supplementary Figure 1** – Schematic representation of the main pre-processing steps for the rsfMRI data
and key subsequent analyses

### 3. Information on quality controls

Quality controls were run to ensure that there was no significant relationship between FD and FC. This was achieved by: (a) plotting a scatterplot of mean FC vs. mean FD with values for each subject; (b) plotting the correlation between edgewise FC and mean FD vs. Euclidean distance between the centroids of all ROIs. This latter plot tells us whether motion affects FC at each edge across different Euclidean distances, i.e. whether motion affects FC in a distance-dependent way (Satterthwaite et al., 2012). This “Satterthwaite” plot (motion correlation plot) was then re-run after regressing-out mean FD from the correlation values (here the intercepts were re-added to the residuals to preserve the variability in FC across edges), achieving gradient=0 and y-intercept=0, and thus ensuring that at the edge-level there was no residual relationship between each subject’s motion and FC. See **Supplementary Figure 2** below for a before and after depiction of this example in (A) the PPMI Cohort and (B) the ICICLE-PD Cohort.

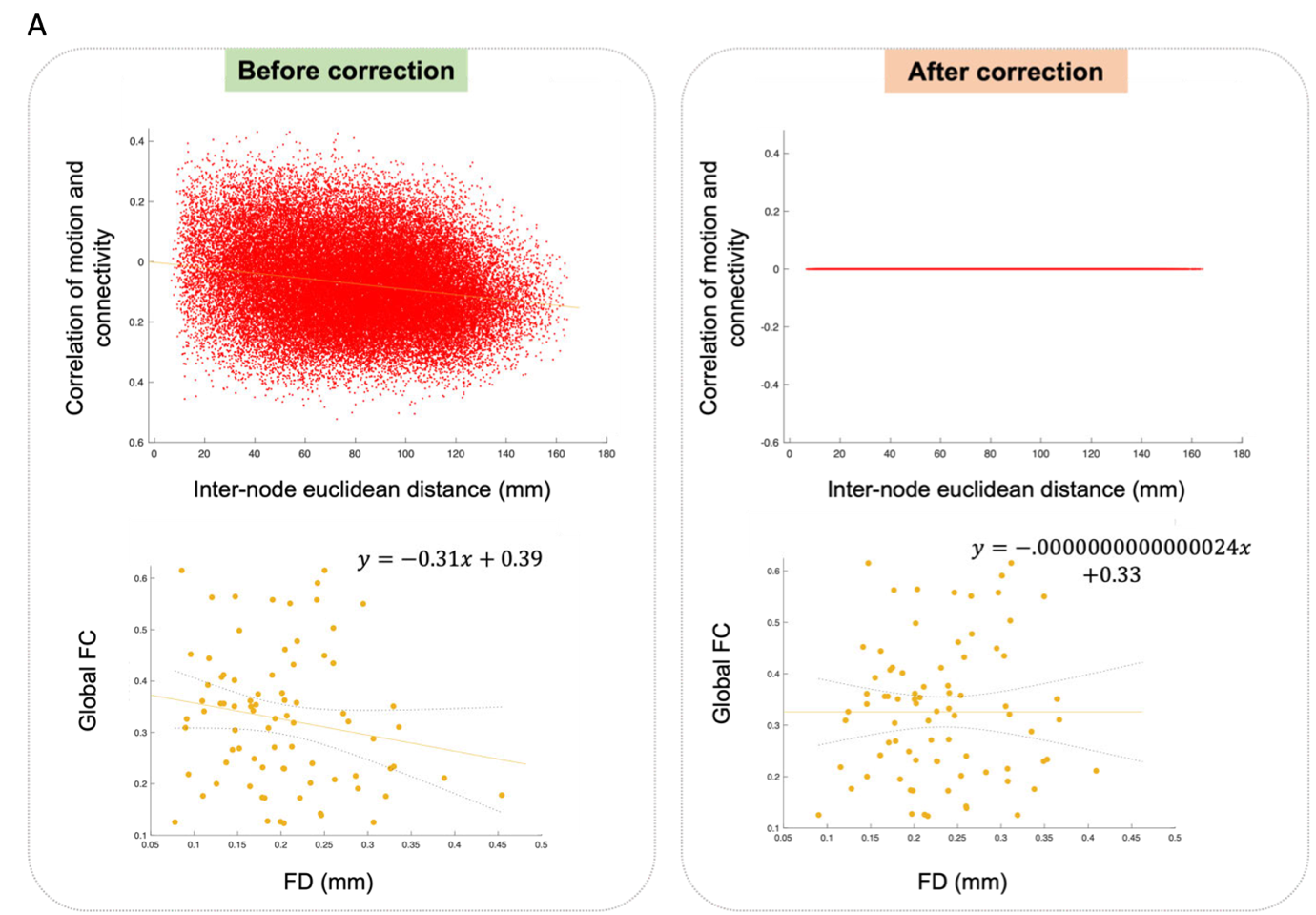

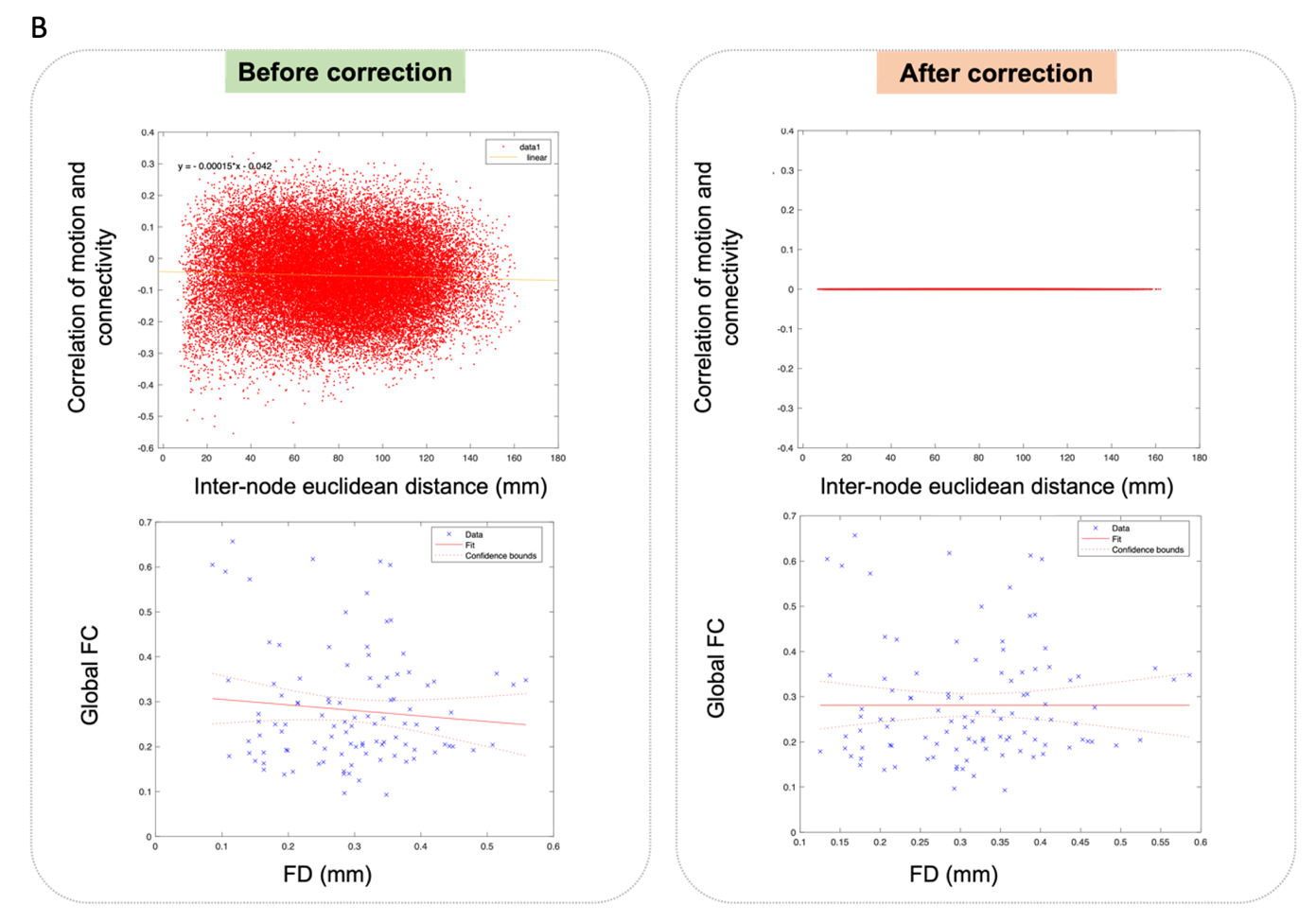

**Supplementary Figure 2 -** Plots of rsfMRI data in the PPMI cohort before and after regressing out FD. The top row shows a “Satterthwaite” plot with the correlation between edgewise FC and mean FD vs. Euclidean distance between the centroids of all ROIs. Bottom row shows scatterplots of mean FC vs. mean FD with values for each subject. (A) PPMI Cohort; (B) ICICLE-PD Cohort.

### 4. Additional information about PPMI and ICICLE-PD Cohorts demographics and comparisons

**Inclusion and Exclusion Criteria for PPMI**

The PPMI cohort included patients diagnosed with idiopathic Parkinson's based on a positive Dopamine Transporter (DaT) scan, as confirmed by expert clinicians using established criteria (Marek et al., 2011). All diagnoses were the provided by the data curators for PPMI  and made by expert clinicians according to established criteria(Marek et al., 2011). Patients with a history of psychotic disorders were excluded. Data was also available for age-matched healthy controls (HC). Exclusion criteria for healthy controls included presence of neurological disorder and having a first degree relative with PD.

**Inclusion and Exclusion Criteria for ICICLE-PD**

Patients with newly diagnosed PD (meeting the UK Brain Bank criteria(Hughes et al., 1992)) were recruited from Newcastle/Gateshead, UK. Ethical approval was provided by the Newcastle and North Tyneside Research Ethics Committee, and written, informed consent was obtained from all participants. Participants were included if they had a diagnosis of idiopathic Parkinson’s and sufficient English for the assessment purposes. Exclusion criteria included having significant cognitive impairment at presentation (Mini Mental State Examination <24), a diagnosis of dementia or atypical Parkinsonism.

**Supplementary Table 1** - Demographics for the full ICICLE-PD cohort with unbalanced groups (*N*=104)

|  | Initial ICICLE Cohort with unbalanced groups | | | |  |
| --- | --- | --- | --- | --- | --- |
| **Variable** | PDNOVH N=55 | | PDVH N=49 | |  |
|  | **Mean** | **SD** | **Mean** | **SD** | **p-val** |
| **Age (y)** | 65.2 | 11.3 | 65.7 | 9.89 | 0.65 |
| **Gender (Females/Males)** | 23/32 | | 11/38 | | 0.03* |
| **MDS-UPDRS III** | 23.9 | 11.4 | 29.1 | 10.5 | 0.01* |
| **Hoehn & Yahr** | 1.89 | 0.59 | 2.02 | 0.69 | 0.32 |
| **PD Duration (y)** | 0.46 | 0.39 | 0.55 | 0.42 | 0.22 |
| **LEDD (mg/day)** | 164.3 | 99.4 | 188 | 170 | 0.68 |
| **Education (y)** | 13.71 | 4.05 | 12.27 | 3.78 | 0.07 |
| **MoCA Total** | 26.2 | 3.16 | 24.91 | 3.46 | 0.03* |
| **Hallucination**  **severity** | - | | 4.05 | 3.61 | - |

Significant p-values <0.05 indicated with *

**Further comparisons between ICICLE and PPMI**

In addition to the summary statistics showing comparison between the PDVH and the PD NOVH patients within each of the cohorts and samples (see **Supplementary Table 1** above and **Table 1** of the main text) the following comparisons across cohorts were also run.

When looking at how ICICLE cohort compared to the PPMI cohort, we ran extra analyses to compare first (A) all PD patients from PPMI to all PD patients in the entire ICICLE cohort (N=104) with Kruskal-Wallis tests across the four groups (PDVH PPMI, PDNOVH PPMI, PDVH ICICLE, PDNOVH ICICLE) and eventual post-hoc tests; and (B) all PPMI compared to the groups in **Sample 1** from ICICLE (i.e., where the ICICLE groups were matched with each other but not with the PPMI cohort); and (C) all PPMI compared to the groups in **Sample 2** from ICICLE (i.e., where all groups were matched within and across cohorts).

For comparison (A) we found that the patients across the two full cohorts and in the different groups were significantly different in terms of MDS-UPDRS-III (p=0.001), years of PD duration (p<0.001), LEDD (p<0.001), years of education (p<0.001) and MoCA total (p<0.001). Post-hoc Dunn's Test with Bonferroni correction showed that the PDVH ICICLE group was significantly different in UPDRS-III from both PDNOVH ICICLE (as was also shown in Table 1) and from the PDNOVH PPMI group (p<0.001); for PD duration both PDVH and PDNOVH groups in ICICLE were significantly different from PDVH PPMI and PDNOVH PPMI (p<0.001), and the same was true when comparing the two cohort in terms of LEDD (all p-val <0.001). Looking at years of education, the NOVH PPMI group was significantly different from both PDVH ICICLE and PDNOVH ICICLE groups (p=0.001 and p=0.01, respectively); finally, as also shown in Table 1, there was a significant difference in MoCA between the two ICICLE groups, in addition to a significant difference between the NOVH PPMI and PDVH ICICLE group (p=0.0003).

For comparison (B) the patients from PPMI and from ICICLE **Sample 1** differed in terms of years of PD duration (p<0.001), LEDD (p<0.001), UPDRS-III (p<0.002), years of education (p<0.001) and MoCA score (p<0.001). Post-hoc Dunn's Test with Bonferroni correction showed that both ICICLE groups had significantly shorter PD duration compared to the PPMI ones (p<0.001) and lower LEDD (p<0.001). PDNOVH patients in the ICICLE cohort also had significantly fewer years of education than their counterparts in the PPMI groups (p<0.05), with the PDVH ICICLE showing this difference at trend significance (p=0.01 but became p=0.07 after multiple comparison correction). Finally, the ICICLE NOVH group also showed significantly lower MoCA score than PDNOVH patients in PPMI (p<0.05).

For comparison (C) the patients from PPMI and from ICICLE **Sample 2** only differed in terms of years of PD duration (p<0.001) and LEDD (p<0.001).

### 5. Detailed NEVHI Results

This section provides a comprehensive breakdown of the responses to the North East Visual Hallucination Interview (NEVHI) Part A screening questions for both Sample 1 and Sample 2 of the ICICLE-PD cohort.

### NEVHI Part A Screening Questions

- 1.1 "Do you feel like your eyes ever play tricks on you? Have you ever seen something (or things) that other people could not see?"
- 1.2 "Have you ever looked at an object or pattern and something else suddenly appeared or disappeared?"
- 1.3 "Have you ever had the feeling of the presence of somebody or something?"
- 1.4 "Have you ever seen a passing shadow in the corner of your eye?"
- 1.5 "Have you ever had other visual experiences?"
- 1.6 "Have you experienced seeing dots, flashes, patterns of light or similar that were not there?"

### Results

#### Sample 1 (n=48) - 33.33% reported multiple types of hallucinations

| **Type of Hallucinatory Experience** | **Number of Participants** | **Percentage** |
| --- | --- | --- |
| 1.1 Complex visual hallucinations | 7 | 14.58% |
| 1.2 Illusions | 5 | 10.42% |
| 1.3 Feeling of presence | 10 | 20.83% |
| 1.4 Passage of Shadow | 18 | 37.50% |
| 1.5 Other visual symptoms | 14 | 29.17% |
| 1.6 Simple visual hallucinations | 11 | 22.92% |

#### Sample 2 (n=25) - 52.00% reported multiple types of hallucinations

| **Type of Hallucinatory Experience** | **Number of Participants** | **Percentage** |
| --- | --- | --- |
| 1.1 Complex visual hallucinations | 6 | 24.00% |
| 1.2 Illusions | 4 | 16.00% |
| 1.3 Feeling of presence | 9 | 36.00% |
| 1.4 Passage of Shadow | 12 | 48.00% |
| 1.5 Other visual symptoms | 7 | 28.00% |
| 1.6 Simple visual hallucinations | 9 | 36.00% |

### 6. Details on the calculation of hallucination severity in ICICLE-PD

If a patient experienced more than one type of hallucinations, the composite severity score for each type was summed to obtain an overall severity score.

- Duration of each hallucinatory episodes measured in NEVHI part A, measured on a 4-point scale: 1 = “less than 5 min”, 2 = “5 min to 2 h”, 3 = “more than 2 h but less than all the time”, 4 = “all the time”.
- Frequency of the hallucinatory episodes, measured on a 4-point scale for each of the types of hallucinations experiences: 0 = “Never”, 1 = “Less than once a week”, 2= “1-6 times a week”, 3 = “Daily”. The total frequency was calculated by adding together the scores for each hallucination experience assessed in the NEVHI part A.
- Severity factors: complex hallucinations=4, illusions=3, passage hallucination=1, feeling of presence= 1, simple hallucinations and others = 1.

These measures were based on questions from the NEVHI and combined across the patient- and caregiver-version of the questionnaire (Mosimann et al., 2008).

### 7. Summary table for Linear models looking at group differences in Yeo networks and von Economo classes

**Supplementary Table 2** - Summary of analyses looking at the effect of group on FC in each Yeo network and vonEconomo class. Significance level <0.05 shown with * or with bold font.

| Class/network (IV) | Estimate | Confidence Interval | Statistics Value | Overall model R^2^ / R^2^ adjusted | P-vals group effect (uncorrected) | P-vals for group effect (FDR corrected) | P-value group contrast |
| --- | --- | --- | --- | --- | --- | --- | --- |
| Von Economo 1 Primary motor | -0.02 | -0.04 – 0.00 | -2.35 | 0.111 / 0.073 | **0.020*** | 0.070 | - |
| Von Economo 2 Association 1 | -0.02 | -0.04 – 0.00 | -2.47 | 0.107 / 0.069 | **0.015*** | 0.070 | - |
| Von Economo 3 Association 2 | -0.02 | -0.04 – 0.00 | -1.98 | 0.080 / 0.041 | 0.051 | 0.088 | - |
| Von Economo 4 Secondary Sensory | -0.01 | -0.03 – 0.01 | -1.11 | 0.084 / 0.045 | 0.270 | 0.270 | - |
| Von Economo 5 Primary sensory | -0.01 | -0.03 – 0.01 | -1.44 | 0.120 / 0.083 | 0.151 | 0.210 | - |
| Von Economo 6 Limbic | -0.02 | -0.04 – 0.00 | -2.04 | 0.099 / 0.060 | **0.043*** | 0.088 | - |
| Von Economo 7 Insular | -0.01 | -0.04 – 0.01 | -1.14 | 0.053 / 0.013 | 0.255 | 0.270 | - |
| Yeo 1  Visual | -0.01 | -0.04 – 0.01 | -1.369 | 0.102 / 0.064 | 0.174 | 0.202 | - |
| Yeo 2  Somatomotor | -0.02 | -0.04 – 0.00 | -2.24 | 0.129 / 0.092 | **0.027*** | 0.065 | - |
| Yeo 3  Dorsal Attention | -0.01 | -0.03 – 0.01 | -1.04 | 0.056 / 0.016 | 0.290 | 0.290 | - |
| Yeo 4  Ventral Attention | -0.02 | 0.04 – 0.00 | -2.14 | 0.085 / 0.046 | **0.034*** | 0.065 | - |
| Yeo 5 Limbic | -0.03 | -0.05 – -0.02 | -4.49 | 0.212 / 0.179 | **<0.001*** | **0.007*** | **<0.001^a^, <0.001^b^**, 0.344^c^ |
| Yeo 6 Frontoparietal | -0.02 | -0.04 – 0.00 | -2.11 | 0.086 / 0.047 | **0.037*** | 0.065 | - |
| Yeo 7 DMN | -0.01 | -0.04 – 0.00 | -1.80 | 0.077 / 0.038 | 0.073 | 0.101 | - |

a- HC vs PDNOVH; b- HC vs PDVH; c- PDNOVH vs PDVH.

### 8. Supplementary details for the different NBS analyses

**Main PPMI NBS network**

**Supplementary Table 3**- Significant connections between node pairs in the NBS subnetwork of reduced FC in patients with PDVH. Pairs are listed in descending order according to their t-statistics value.

| Edge | Node Pair [i , j] Anatomical region | | Functional  Yeo network [ i , j ] | | T-stat value |
| --- | --- | --- | --- | --- | --- |
| **#** | **Node_i** | **Node_j** | **Node_i** | **Node_j** |  |
| 19 | lh_precentral_part8 | rh_superiorfrontal_part12 | Dorsal attention | DMN | 4.59 |
| 20 | lh_precentral_part8 | rh_superiorfrontal_part13 | Dorsal attention | Frontoparietal | 4.59 |
| 15 | lh_precentral_part8 | rh_superiorfrontal_part11 | Dorsal attention | DMN | 4.40 |
| 18 | rh_postcentral_part8 | rh_superiorfrontal_part11 | Somatomotor | DMN | 4.32 |
| 16 | lh_superiorparietal_part1 | rh_superiorfrontal_part11 | Somatomotor | DMN | 4.23 |
| 11 | lh_precentral_part6 | rh_precuneus_part3 | Somatomotor | Dorsal attention | 4.22 |
| 23 | rh_superiorfrontal_part11 | rh_superiorparietal_part9 | DMN | Dorsal attention | 4.20 |
| 3 | lh_paracentral_part3 | lh_superiorfrontal_part11 | Somatomotor | DMN | 4.15 |
| 10 | lh_paracentral_part2 | rh_precuneus_part3 | Somatomotor | Dorsal attention | 4.11 |
| 1 | lh_precentral_part8 | lh_superiorfrontal_part9 | Dorsal attention | DMN | 4.05 |
| 2 | lh_paracentral_part1 | lh_superiorfrontal_part11 | Somatomotor | DMN | 4.04 |
| 4 | lh_superiorfrontal_part13 | rh_paracentral_part2 | Frontoparietal | Somatomotor | 4.03 |
| 8 | lh_superiorfrontal_part10 | rh_precentral_part8 | Ventral att. | Somatomotor | 3.94 |
| 7 | lh_superiorfrontal_part11 | rh_precentral_part6 | DMN | Somatomotor | 3.89 |
| 21 | lh_superiorparietal_part1 | rh_superiorfrontal_part13 | Somatomotor | Frontoparietal | 3.89 |
| 6 | lh_superiorfrontal_part11 | rh_paracentral_part3 | DMN | Somatomotor | 3.88 |
| 5 | lh_superiorparietal_part1 | rh_paracentral_part2 | Somatomotor | Somatomotor | 3.87 |
| 9 | lh_superiorfrontal_part13 | rh_precentral_part8 | Frontoparietal | Somatomotor | 3.87 |
| 12 | lh_precentral_part8 | rh_precuneus_part3 | Dorsal attention | Dorsal attention | 3.85 |
| 13 | lh_rostralmiddlefrontal_part9 | rh_precuneus_part3 | Frontoparietal | Dorsal attention | 3.84 |
| 14 | lh_paracentral_part3 | rh_superiorfrontal_part11 | Somatomotor | DMN | 3.84 |
| 17 | rh_paracentral_part2 | rh_superiorfrontal_part11 | Somatomotor | DMN | 3.82 |
| 22 | rh_superiorfrontal_part7 | rh_superiorparietal_part9 | Ventral att. | Dorsal attention | 3.81 |

Importantly, the NBS subnetwork found in the PPMI cohort remained relatively unchanged after more careful matching of patients according to disease severity (UPDRS-III) – although there was no statistical difference in this variable to begin with. This was tested with a sensitivity analysis with matched PPMI patients according to disease severity and demographic variables, leading to PDVH N=25 and PDNOVH N=25. The same NBS parameters were used, namely t-threshold=3.8 and 5000 permutations. See Supplementary Figure 3 below.

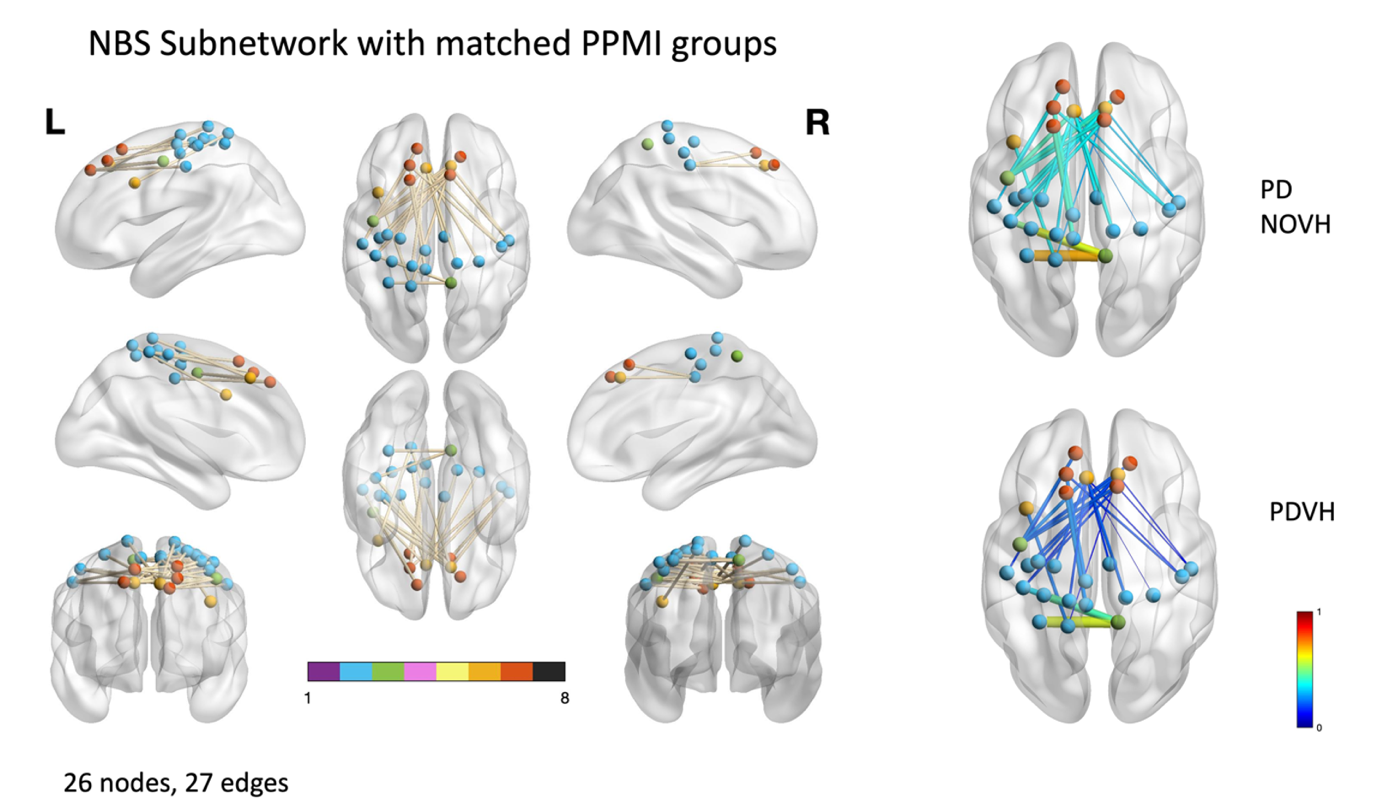

**Supplementary Figure 3** – NBS Subnetwork in PPMI cohort after more careful UPDRS-III matching (PDVH N=25, PDNOVH N=25).

### 9. ICICLE-PD Cohort - further analyses

No NBS subnetwork at all was found in **Sample 1.**

Below is the NBS subnetwork found in **Sample 2** after matching patients within and across cohorts as detailed in the main text:

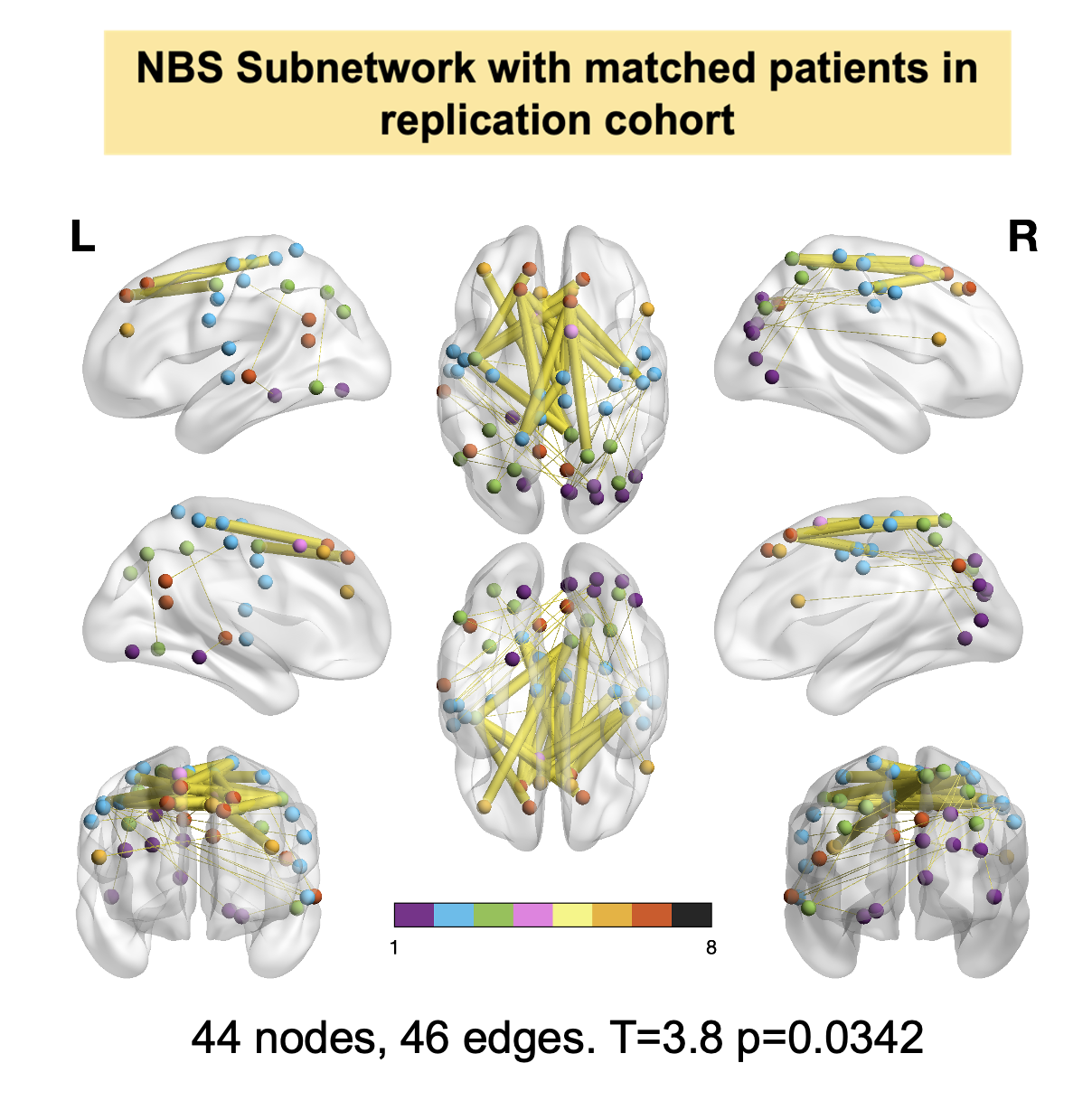

**Supplementary Figure 4-** Replication of the NBS subnetwork in ICICLE-PD dataset after patient matching. Here the results indicate an NBS subnetwork with patients in replication cohort (VH vs NOVH) where PDVH have lower FC than NOVH. The same edges as the original NBS subnetwork from PPMI have been coloured in yellow. Nodes are colour-coded according to their mapping to the 7 canonical functional networks by Yeo et al. (2011).

**Supplementary Table 4** - Significant connections between node pairs in the NBS subnetwork of reduced FC in patients with PDVH in the ICICLE-PD Cohort.

| Edge | Node Pair [i , j] Anatomical region | | T-stat value |
| --- | --- | --- | --- |
| **#** | **Node_i** | **Node_j** |  |
| 1 | lh_paracentral_part2 | lh_precuneus_part7 | 3.87 |
| 2 | lh_precentral_part8 | lh_superiorfrontal_part9 | 5.16 |
| 3 | lh_paracentral_part1 | lh_superiorfrontal_part11 | 4.49 |
| 4 | lh_paracentral_part3 | lh_superiorfrontal_part11 | 6.38 |
| 5 | lh_inferiortemporal_part2 | lh_superiorparietal_part9 | 4.25 |
| 6 | lh_parahippocampal_part2 | lh_superiortemporal_part5 | 4.25 |
| 7 | lh_superiorparietal_part4 | lh_superiortemporal_part5 | 4.68 |
| 8 | lh_postcentral_part2 | rh_cuneus_part1 | 4.88 |
| 9 | lh_superiortemporal_part5 | rh_cuneus_part1 | 6.21 |
| 10 | lh_superiortemporal_part6 | rh_cuneus_part1 | 6.2 |
| 11 | lh_superiorfrontal_part13 | rh_paracentral_part2 | 4.3 |
| 12 | lh_superiorparietal_part1 | rh_paracentral_part2 | 5.27 |
| 13 | lh_superiorfrontal_part11 | rh_paracentral_part3 | 4.87 |
| 14 | lh_inferiorparietal_part4 | rh_parsopercularis_part2 | 4.26 |
| 15 | rh_cuneus_part1 | rh_parsopercularis_part2 | 3.87 |
| 16 | rh_cuneus_part1 | rh_postcentral_part3 | 5.05 |
| 17 | rh_lateraloccipital_part2 | rh_postcentral_part3 | 4.42 |
| 18 | rh_pericalcarine_part3 | rh_postcentral_part3 | 4.2 |
| 19 | lh_inferiortemporal_part2 | rh_postcentral_part8 | 5.7 |
| 20 | lh_lingual_part6 | rh_postcentral_part8 | 3.87 |
| 21 | lh_precentral_part8 | rh_postcentral_part8 | 3.87 |
| 22 | rh_inferiorparietal_part2 | rh_postcentral_part8 | 5.35 |
| 23 | lh_superiorfrontal_part11 | rh_precentral_part6 | 4.87 |
| 24 | lh_paracentral_part2 | rh_precuneus_part3 | 6.35 |
| 25 | lh_precentral_part6 | rh_precuneus_part3 | 4.47 |
| 26 | lh_precentral_part8 | rh_precuneus_part3 | 7.67 |
| 27 | lh_paracentral_part2 | rh_precuneus_part6 | 4.27 |
| 28 | lh_inferiorparietal_part4 | rh_superiorfrontal_part11 | 4.65 |
| 29 | lh_paracentral_part3 | rh_superiorfrontal_part11 | 4.94 |
| 30 | lh_superiorparietal_part1 | rh_superiorfrontal_part11 | 4.07 |
| 31 | rh_inferiorparietal_part4 | rh_superiorfrontal_part11 | 5.18 |
| 32 | rh_paracentral_part2 | rh_superiorfrontal_part11 | 4.89 |
| 33 | rh_postcentral_part8 | rh_superiorfrontal_part11 | 4.7 |
| 34 | lh_precentral_part8 | rh_superiorfrontal_part12 | 5.61 |
| 35 | lh_precentral_part8 | rh_superiorfrontal_part13 | 3.95 |
| 36 | lh_superiortemporal_part6 | rh_superiorparietal_part1 | 5.69 |
| 37 | lh_inferiorparietal_part3 | rh_superiorparietal_part4 | 3.88 |
| 38 | lh_postcentral_part2 | rh_superiorparietal_part4 | 4.11 |
| 39 | lh_precentral_part7 | rh_superiorparietal_part4 | 4.58 |
| 40 | lh_precentral_part9 | rh_superiorparietal_part4 | 5.07 |
| 41 | rh_postcentral_part6 | rh_superiorparietal_part4 | 3.95 |
| 42 | rh_postcentral_part8 | rh_superiorparietal_part4 | 4 |
| 43 | rh_precentral_part7 | rh_superiorparietal_part4 | 4.09 |
| 44 | rh_superiorfrontal_part11 | rh_superiorparietal_part9 | 4.42 |
| 45 | rh_cuneus_part1 | rh_superiorparietal_part10 | 4.45 |
| 46 | rh_pericalcarine_part3 | rh_superiorparietal_part10 | 5.01 |

10. **Linear regression models between NBS FC at rsfMRI timepoint (baseline) and follow-up scores after adjusting for outcome scores at scanning time.**

**Supplementary Table 5 – Linear regression models between NBS FC at rsfMRI timepoint (baseline) and follow-up scores after adjusting for outcome scores at scanning time.**

| Group | Score at follow-up  4 years (± 1 year)  (DVs) | Estimate | Confidence Interval | Statistics  Value | P-val for NBS FC |
| --- | --- | --- | --- | --- | --- |
| VH | MoCA Tot Score | 5.23 | -5.18 – 15.63 | 1.042 | 0.309 |
| NOVH | MoCA Tot Score | 4.08 | -1.01 – 9.18 | 1.607 | 0.114 |
| VH | Attention | 1.05 | -6.73 – 8.83 | 0.281 | 0.781 |
| NOVH | Attention | 0.02 | -6.87 – 6.92 | 0.007 | 0.994 |
| VH | RBD | 0.12 | -9.41 – 9.66 | 0.027 | 0.979 |
| NOVH | RBD | -3.54 | -10.32 – 3.25 | -1.049 | 0.300 |
| VH | MDS-UPDRS III | -27.81 | -72.18 – 16.56 | -1.300 | 0.207 |
| NOVH | MDS-UPDRS III | 4.45 | -23.91 – 32.81 | 0.315 | 0.754 |

11. **Information on software used and links to open-access datasets**

| Resource | Citation | Link |
| --- | --- | --- |
| FreeSurfer | Fischl (2012) | <http://surfer.nmr.mgh.harvard.edu> |
| FSL | FMRIB Analysis Group | <https://fsl.fmrib.ox.ac.uk/fsl/fslwiki/> |
| AFNI | Cox (1996) | <https://afni.nimh.nih.gov/> |
| SPM | Wellcome Trust Centre for Neuroimaging (UCL) | [www.fil.ion.ucl.ac.uk/spm](http://www.fil.ion.ucl.ac.uk/spm) |
| R | R Development Core Team, (2008) | <https://www.r-project.org/> |
| RStudio | RStudio | <https://www.rstudio.com/> |
| Python | Python Software Foundation | <https://www.python.org/> |
| MATLAB | MathWorks | [www.mathworks.com](http://www.mathworks.com) |
| BrainNet Viewer | Xia et al., (2013) | <https://www.nitrc.org/projects/bv/> |
| Brain Connectivity Toolbox | Rubinov M, Sporns O (2010) | <https://sites.google.com/site/bctnet/> |
| Brains for publication | Whitaker, Notter, & Morgan (2017) | <https://github.com/WhitakerLab/BrainsForPublication> |
| PPMI Dataset | Parkinson’s Progression Markers Initiative | <https://www.ppmi-info.org/> |
| ICICLE-PD Dataset | Burn, et al., (2017) | <http://bam-ncl.co.uk/our-work/studies/icicle-pd/> |

**Supplementary Material References**

Desikan, R. S., Ségonne, F., Fischl, B., Quinn, B. T., Dickerson, B. C., Blacker, D., Buckner, R. L., Dale, A. M., Maguire, R. P., Hyman, B. T., Albert, M. S., & Killiany, R. J. (2006). An automated labeling system for subdividing the human cerebral cortex on MRI scans into gyral based regions of interest. *NeuroImage*, *31*(3), 968–980. https://doi.org/10.1016/j.neuroimage.2006.01.021

Fischl, B., Sereno, M. I., & Dale, A. M. (1999). Cortical surface-based analysis. II: Inflation, flattening, and a surface-based coordinate system. *NeuroImage*, *9*(2), 195–207. https://doi.org/10.1006/nimg.1998.0396

Glasser, M. F., Coalson, T. S., Robinson, E. C., Hacker, C. D., Harwell, J., Yacoub, E., Ugurbil, K., Andersson, J., Beckmann, C. F., Jenkinson, M., Smith, S. M., & Van Essen, D. C. (2016). A multi-modal parcellation of human cerebral cortex. *Nature*, *536*(7615), 171–178. https://doi.org/10.1038/nature18933

Mosimann, U. P., Collerton, D., Dudley, R., Meyer, T. D., Graham, G., Dean, J. L., Bearn, D., Killen, A., Dickinson, L., Clarke, M. P., & McKeith, I. G. (2008). A semi-structured interview to assess visual hallucinations in older people. *International Journal of Geriatric Psychiatry*, *23*(7), 712–718. https://doi.org/10.1002/gps.1965

Patel, A. X., Kundu, P., Rubinov, M., Jones, P. S., Vértes, P. E., Ersche, K. D., Suckling, J., & Bullmore, E. T. (2014). A wavelet method for modeling and despiking motion artifacts from resting-state fMRI time series. *NeuroImage*, *95*, 287–304. https://doi.org/10.1016/j.neuroimage.2014.03.012

Romero-Garcia, R., Atienza, M., Clemmensen, L. H., & Cantero, J. L. (2012). Effects of network resolution on topological properties of human neocortex. *NeuroImage*, *59*(4), 3522–3532. https://doi.org/10.1016/j.neuroimage.2011.10.086

Satterthwaite, T. D., Wolf, D. H., Loughead, J., Ruparel, K., Elliott, M. A., Hakonarson, H., Gur, R. C., & Gur, R. E. (2012). Impact of in-scanner head motion on multiple measures of functional connectivity: Relevance for studies of neurodevelopment in youth. *NeuroImage*, *60*(1), 623–632. https://doi.org/10.1016/j.neuroimage.2011.12.063

Váša, F., Bullmore, E. T., & Patel, A. X. (2018). Probabilistic thresholding of functional connectomes: Application to schizophrenia. *NeuroImage*, *172*, 326–340. https://doi.org/10.1016/j.neuroimage.2017.12.043
